# Do nutritional interventions before or during pregnancy affect placental phenotype? Findings from a systematic review of human clinical trials

**DOI:** 10.1101/2024.05.15.24307442

**Authors:** V Bonnell, M White, KL Connor

## Abstract

**Background:** Maternal nutritional interventions aim to address nutrient deficiencies in pregnancy, a leading cause of maternal and neonatal morbidity and mortality worldwide. How these interventions influence the placenta, which plays a vital role in fetal growth and nutrient supply, is not well understood. This is a major gap in understanding how such interventions could influence pregnancy outcomes and fetal health. We hypothesised that nutritional interventions influence placental phenotype, and that these placental changes relate to how successful, or not, the intervention is in improving pregnancy outcomes.

**Methods:** We conducted a systematic review and followed PRISMA-2020 reporting guidelines. Articles were retrieved from PubMed, Clinicaltrials.gov, and ICTRP-WHO using pre-defined search terms and screened by two reviewers using a 3-level process. Inclusion criteria considered articles published from January 2001-September 2021 that reported on clinical trials in humans, which administered a maternal nutritional intervention during the periconceptional or pregnancy period and reported on placental phenotype (shape and form, function or placental disorders).

**Findings:** Fifty-three eligible articles reported on (multiple) micronutrient- (n=33 studies), lipid- (n=11), protein- (n=2), and diet-/lifestyle-based (n=8) interventions. Of the micronutrient-based interventions, 16 (48%) associated with altered placental function, namely altered nutrient transport/metabolism (n=9). Nine (82%) of the lipid-based interventions associated with altered placental phenotype, including elevated placental fatty acid levels (n=5), altered nutrient transport/metabolism gene expression (n=4), and decreased inflammatory biomarkers (n=2). Of the protein-based interventions, two (66%) associated with altered placental phenotype, including increased placental efficiency (n=1) or decreased preeclampsia risk (n=1). Three (38%) of diet and lifestyle-based interventions associated with placental changes, namely placental gene expression (n=1) and disease (n=2). In studies with data on maternal (n=30) or offspring (n=20) outcomes, interventions that influenced placental phenotype were more likely to have also associated with improved maternal outcomes (11/15 [73%]) and offspring birth outcomes (6/11 [54%]), compared to interventions that did not associate with placental changes (2/15 [13%] and 1/9 [11%], respectively).

**Conclusions:** Periconceptional and prenatal nutritional interventions to improve maternal/pregnancy health associate with altered placental development and function. These placental adaptations likely benefit the pregnancy and improve offspring outcomes. Understanding the placenta’s role in the success of interventions to combat nutrient deficiencies is critical for improving interventions and reducing maternal and neonatal morbidity and mortality globally.

## Introduction

Maternal and child undernutrition contribute to around 50% of global deaths in children under five (1). Nutrient deficiencies in mothers can worsen during pregnancy due to increased nutritional demands needed to support both the mother and developing fetus (1, 2). The periconceptional and perinatal periods are critical windows for offspring development, and nutritional inadequacies during these times associate with both immediate adverse outcomes for the offspring, such as an increased risk of poor fetal growth, stunting, preterm birth, and mortality (2, 3), and long-term health issues, including increased risk of type 2 diabetes, hypertension, and coronary heart disease later in life (4, 5). For mothers, poor nutritional status during these periods can increase the risk of anaemia, hypertension, miscarriage, or mortality (6). Common nutritional interventions to improve maternal and pregnancy outcomes include daily supplementation with folic acid (7), with and without iron, multiple micronutrients, or calcium (8). However, these interventions have shown inconsistent success in improving pregnancy outcomes (2, 9), in part because the biological mechanisms that determine an intervention’s impact are complex and remain poorly understood.

The placenta plays a crucial but often overlooked biological role in how nutritional interventions affect maternal, fetal, and infant health outcomes. As a key organ in pregnancy that develops alongside the embryo/fetus, the placenta plays a vital role in fetal growth, facilitating nutrient transport (glucose, amino acids, lipids, and vitamins and minerals (10, 11)), waste removal, and protection from harmful substances (10). Placental nutrient transport is influenced by nutrient availability and fetal nutrient requirements (12), among other factors, and can adapt to buffer against temporary fluctuations in maternal nutrient status, whether deficiencies or oversupply, to maintain a stable nutritional environment for the developing fetus (10). Importantly, the placenta itself is a developing organ that requires adequate nutritional resources to develop and function properly, and inadequate resources may disrupt optimal placental development and function (13). Nutritional interventions have the potential to increase nutrient availability for the developing placenta, which can in turn influence placental gene expression, angiogenesis, nutrient transport, and activity in inflammatory and oxidative stress pathways, among other functions (14, 15). A healthy placenta also benefits maternal health in the short-term by releasing placental hormones that can alter maternal physiology in preparation for pregnancy and lactation (16), and placental syndromes can have long-term cardiovascular consequences for the mother (17). Therefore, nutritional interventions aimed at improving maternal and fetal health may be associated with changes in the placenta, and understanding these changes is important for determining why these interventions are successful in some contexts but not others.

To-date, reviews on how nutritional interventions influence the placenta have mainly focused on placental-related pregnancy complications, such as preeclampsia (18–21), rather than on the overall placental phenotype. In this systematic review, we synthesised existing knowledge on how direct maternal nutritional interventions in the periconceptional or pregnancy periods impact placental phenotype. We aimed to determine whether: 1. improved maternal and offspring outcomes were more likely to occur with nutritional interventions that reported placental phenotype alterations, and 2. associations between nutritional interventions and placental phenotype differed based on placental sex or study location. We hypothesised that direct maternal nutritional interventions in the periconceptional and pregnancy periods would be associated with altered placental phenotype, and that nutritional interventions that improved maternal and offspring health would more likely associate with beneficial placental adaptations compared to interventions without improvements in maternal or offspring outcomes.

## Methods

Additional details on the methods used can be found in the Supplementary Methods.

### Reporting guidelines

This study adhered to current Preferred Reporting Items for Systematic Reviews and Meta-Analyses (PRISMA-2020)(22) (Figure 1) and Synthesis without meta-analysis (SWiM) (23) frameworks (Supplementary Table S1).

**Figure 1.**
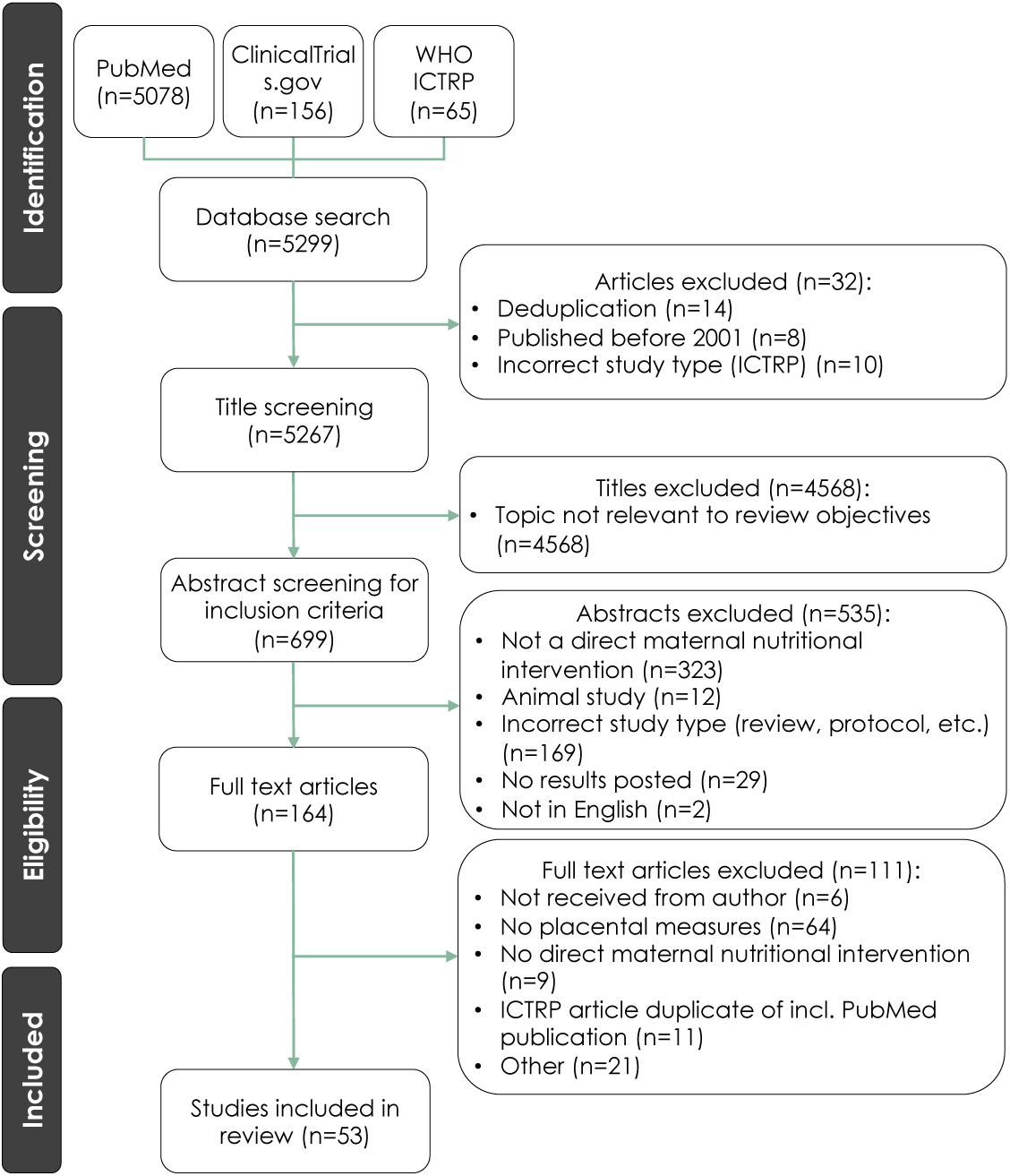
PRISMA flow diagram for article selection.

### Inclusion and exclusion criteria

Eligible study designs included randomised and non-randomised controlled trials in human populations that administered a direct nutritional intervention to the mother during the pre-conception, periconceptional, or pregnancy periods. Inclusion criteria also required that studies were written in English and published between January 2001 and September 2021 (time of search). To increase the translatability of the review findings to the current day context, we limited our review to studies published from 2001 onwards, as this period followed the implementation of folic acid fortification and peri-conceptional supplementation in several countries (24), thereby reducing the potential for including studies where folate deficiency or insufficiency in pregnancy was common and likely to have effects on placental development and function (25).

The population of interest was pregnant people of any age receiving a direct nutritional intervention before or during any stage of pregnancy (Supplementary Figure S1). Interventions were considered direct if they were nutrition-specific and administered directly to the mother (vs. an indirect nutritional intervention, which may seek to address the underlying drivers of undernutrition, and may target household income/food security, health services, water, sanitation, and food production) (26). Studies were required to have included a comparison group that served as a control, and some measure of placental phenotype as an outcome (but not necessarily the primary outcome of that study; Supplementary Figure S1). Placental outcomes were categorized into the following five groups: anthropometry (any physical characteristics related to the placenta), molecular (any alterations to placental genotype and/or gene expression, or other molecular changes), pathology (any findings related to placental histopathology), placental abruption (cases of placental abruption), and placenta-related disease (any disease with placental origins, or that significantly impacts the placenta; Supplementary Table S2).

### Information sources and search terms

PubMed, ClinicalTrials.gov, and the World Health Organization (WHO) International Clinical Trials Registry Platform (ICTRP) were searched to identify peer-reviewed publications using predefined search terms (Supplementary Methods).

### Article search, screening, and data collection

A three-level screening process was performed by two authors (VB and MW; Figure 1). A total of 5299 titles from PubMed (n=5078), ClinicalTrials.gov (n=156), and WHO ICTRP (n=65) were captured from the article search. Following deduplication and title screening (Level 1) and abstract screening (Level 2), 164 records were carried forward for full-text screening (Level 3; Supplementary Methods). After exclusion for not including a placental measure (n=64; including articles reporting on relationships between a nutritional intervention and risk of preeclampsia that did not provide placenta-specific measures), not administering a direct nutritional intervention (n=9), duplicates (n=11), or other reasons (i.e., wrong study type, no trial results available or could not be found (n=21), 53 articles remained and met the full inclusion criteria.

### Data extraction

Data were extracted from each of the 53 articles, including study location, maternal clinical characteristics and demographics, type of pregnancy (singleton or twin), and nutritional intervention details (type [micronutrient, macronutrient, and/or diet- and lifestyle-based], composition, timing, dose, and compliance data) (Supplementary Methods). Data on maternal comorbidities reported in the studies under review were noted and used to inform results interpretations. Reported adverse outcomes were noted and classified into two categories: adverse effects (outcomes that were suspected to be in response to the intervention (27)) and adverse events (outcomes that were not suspected to be in response to the intervention (27)). Data on associations between the nutritional intervention and 1. placental phenotype (primary outcome), and 2. placental sex, and 3. fetal/infant and maternal outcomes (secondary outcomes) were also captured and summarised, and the proportion of studies that did or did not report placental changes or improvements in maternal and offspring outcomes was calculated for each subtype of nutritional intervention (micronutrient, macronutrient, and diet- and lifestyle-based). Study outcomes were considered ‘improved’ if the adverse clinical or physiological condition was prevented or bettered in mothers or offspring. Examples of improved outcomes included a reduced preeclampsia or preterm birth risk, or improved maternal nutritional status or fetal growth.

### Risk of bias assessments

The articles under review were assessed for risk of bias (RoB) using the Cochrane Collaboration’s Tool for Assessing Risk of Bias for randomised studies (n=50; Supplementary Table S3) and the Risk of Bias in Non-randomised Studies of Interventions (ROBINS-I) tool for non-randomised studies (n=3) (28, 29) (Supplementary Methods). RoB assessments were performed independently by two authors (VB and MW). Discrepancies in RoB assessments were resolved through discussion with a third author (KLC). While RoB in interventions was not a primary outcome of this review, assessments of RoB were performed to inform results interpretations.

### Statistical analysis

Data were analysed using JMP 16.0. Relative risk (RR; 95% confidence interval) was calculated using a 2x2 table analysis with p value from Fisher’s exact test (2-tail) to determine whether nutritional interventions that improved maternal or offspring outcomes were more likely to associate with placental changes than interventions that did not improve maternal or offspring outcomes. Due to the high heterogeneity of the interventions administered, placental characteristics measured, and statistics reported across studies, a meta-analysis of intervention effects on placental outcomes was not feasible and results were summarised descriptively.

## Results

### Study and cohort characteristics

The 53 included studies were randomised controlled clinical trials (n=43 [81%]), interventional clinical trials (n=7 [13%]), controlled feeding studies (n=2 [4%]), and intention-to-treat studies (n=1 [2%]). These studies administered micronutrient- (n=33 [62%]), lipid- (n=11 [21%]), protein- (n=3 [6%]) and diet and lifestyle-based (n=8 [15%]) interventions. The included studies reported on cohorts from LICs (n=4 [7.5%]), LMICs (n=12 [23%]), UMICs (n=9 [17%]), and HICs (n=33 [62%]; Figures 2-3). Five studies reported findings from multiple cohorts, which included participants from countries with differing socioeconomic statuses.

**Figure 2.**
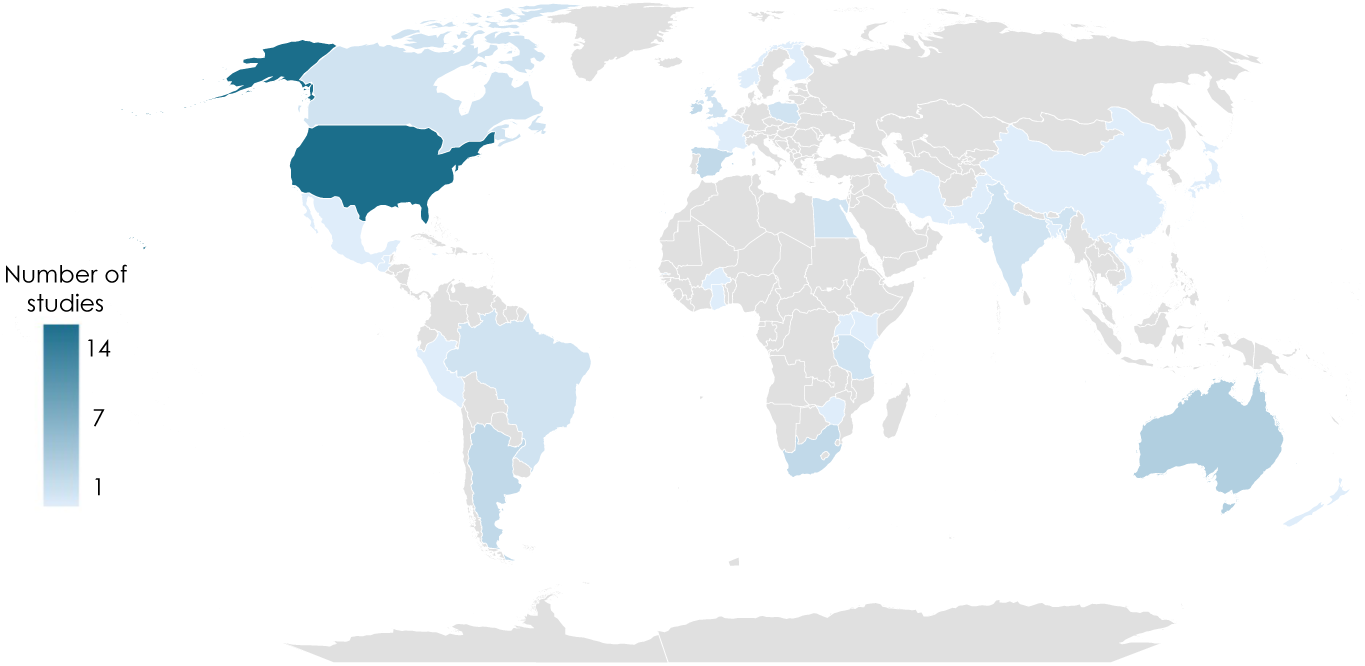
Geographical distribution of the cohorts included in the 53 articles reviewed.

**Figure 3.**
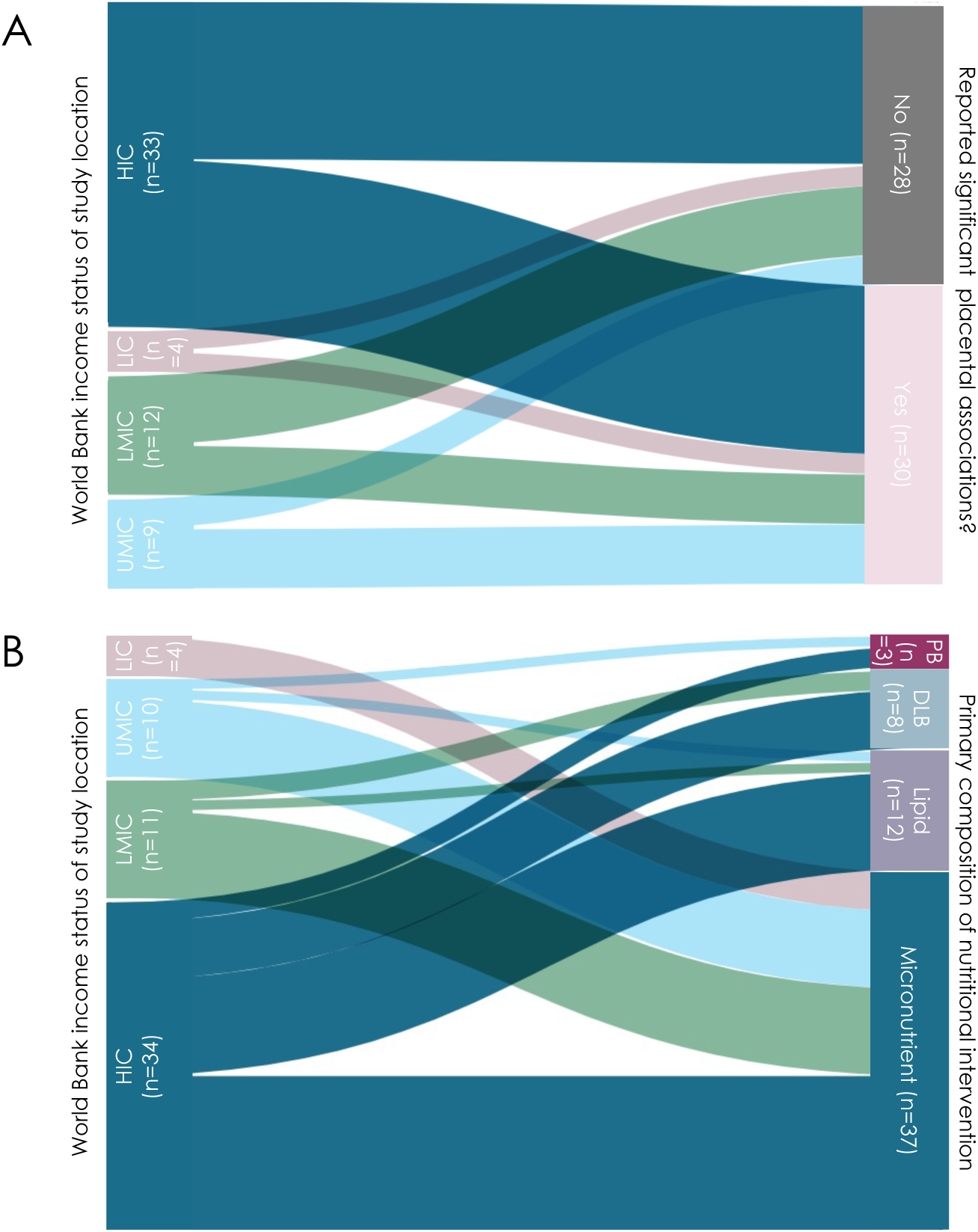
Income status of locations and (A) placental changes and (B) types of nutritional interventions in the studies included in the review. Diagram created on app.rawgraphs.io website. LIC = low-income country. LMIC = low-middle income country. UMIC = upper-middle income country. HIC = high-income country. n = number of studies. DLB = Diet and lifestyle-based. PB = protein-based.

The majority (n=46 [87%]) of studies included only singleton pregnancies, while seven (13%) included both singleton and twin pregnancies. Maternal comorbidities were common in the studies under review (n=33 [62%]) and were mainly hypertensive or metabolic disorders in pregnancy. Further details on the cohort characteristics and adverse effects and events are provided in the Supplementary Results.

Interventions began either peri- (n=3 [6%]) or post-conceptionally (n=50 [94%]; 14 beginning in the first trimester, 28 in the second trimester, and 11 in the third trimester), with the majority continuing until birth (n=52 [98%]), and one that finished after 12 weeks of administration. Key technical terms are defined in Table 1.

**Table 1.** Definitions of key technical terms.

| Term | Definition | Relevance to review objectives |
| --- | --- | --- |
| Amnion-chorio-decidua separation | The separation of a protective membrane surrounding the fetus filled with fluid (amnion) and the location of fetal attachment to the placenta (chorion decidua) | Separation that occurs at the incorrect time (after ~16 weeks' gestation), can increase risk of preterm birth or stillbirth |
| Cerebro-placental ratio | A measure of how blood flows to the fetal brain versus the placenta; derived from the doppler index (a measure of how blood flows) of the middle cerebral artery compared to the umbilical artery | An abnormal (lower) ratio is caused by changes in vascular dilation and placental resistance, and is associated with preterm birth risk, lower birth weight, and fetal distress |
| Chorioamnionitis | Infection and inflammation of the amnion and chorion (fetal membranes); characterised by intra-amniotic neutrophil infiltration | Associated with adverse pregnancy and fetal outcomes, including increased risk of preterm birth |
| Histomorphometry | The quantitative analysis of measurable traits in tissues (i.e., size, area) | Placental histomorphometry can provide insight into placental development and function |
| Histopathology | Atypical results confirmed via microscopic analysis | Positive histopathological finding indicates atypical results (such as the presence of a disease) |
| Hypertension | High blood pressure | Hypertension is associated with increased risk of maternal and fetal mortality, preterm birth, and preeclampsia |
| Placental abruption | Separation of the placenta from the uterus during pregnancy | Can lead to maternal and/or fetal mortality, depending on the stage of pregnancy |
| Preeclampsia | A placental-mediated hypertensive disorder that can occur after 20 weeks of pregnancy | A leading cause of maternal and fetal mortality |
| Premature rupture of membranes | Rupturing of the amniotic sac before labour contractions begin | Can increase risk of maternal and fetal infection and mortality |
| Twin-to-twin transfusion syndrome | A condition where there is an imbalance in blood flow and nutrients between twins that share a single placenta | Can have harmful effects on both fetuses through either lack of or excess blood flow, such as insufficient nutrient intake |

### Risk of bias assessments

All 50 randomised studies under review had low or unclear risk of selection, detection, attrition, and reporting bias (Figure 4). Risk of performance bias was assessed to be high in 5 trials (9%) where blinding was not possible given the nature of the nutritional intervention (Figure 4). Most studies (n=38 [72%]) had a low RoB related to intervention compliance, however, methods for measuring intervention compliance were unclear in over one quarter of the studies under review (n=14 [26%]), and one study was assessed to have a high RoB due to limited participant follow up and no reported compliance measures (Figure 4) (30).

**Figure 4.**
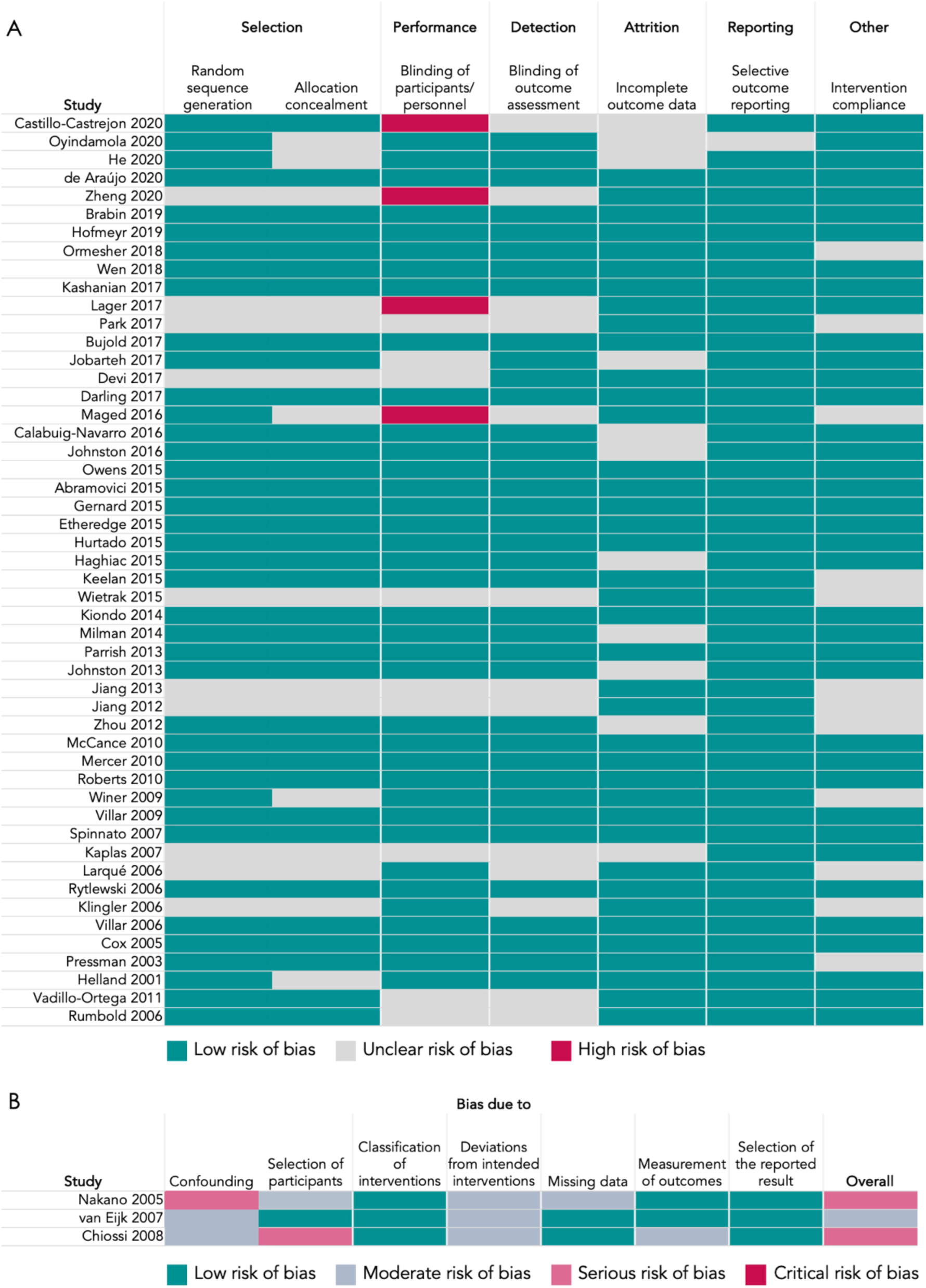
Quality assessments of randomised studies using the Cochrane Collaboration’s tool for Assessing Risk of Bias (A) and non-randomised studies using the ROBINS-I assessment tool (B).

Of the three non-randomised studies under review, one had an overall moderate RoB and two had an overall serious RoB (Figure 5). Domain-specific contributors to a serious RoB were a lack of controlling for confounding variables (30) and methods used for selection of participants into the study (31). All three non-randomised studies had a moderate RoB due to potential deviations from the intended interventions, which was generally due to a lack of recording or reporting on intervention compliance.

**Figure 5.**
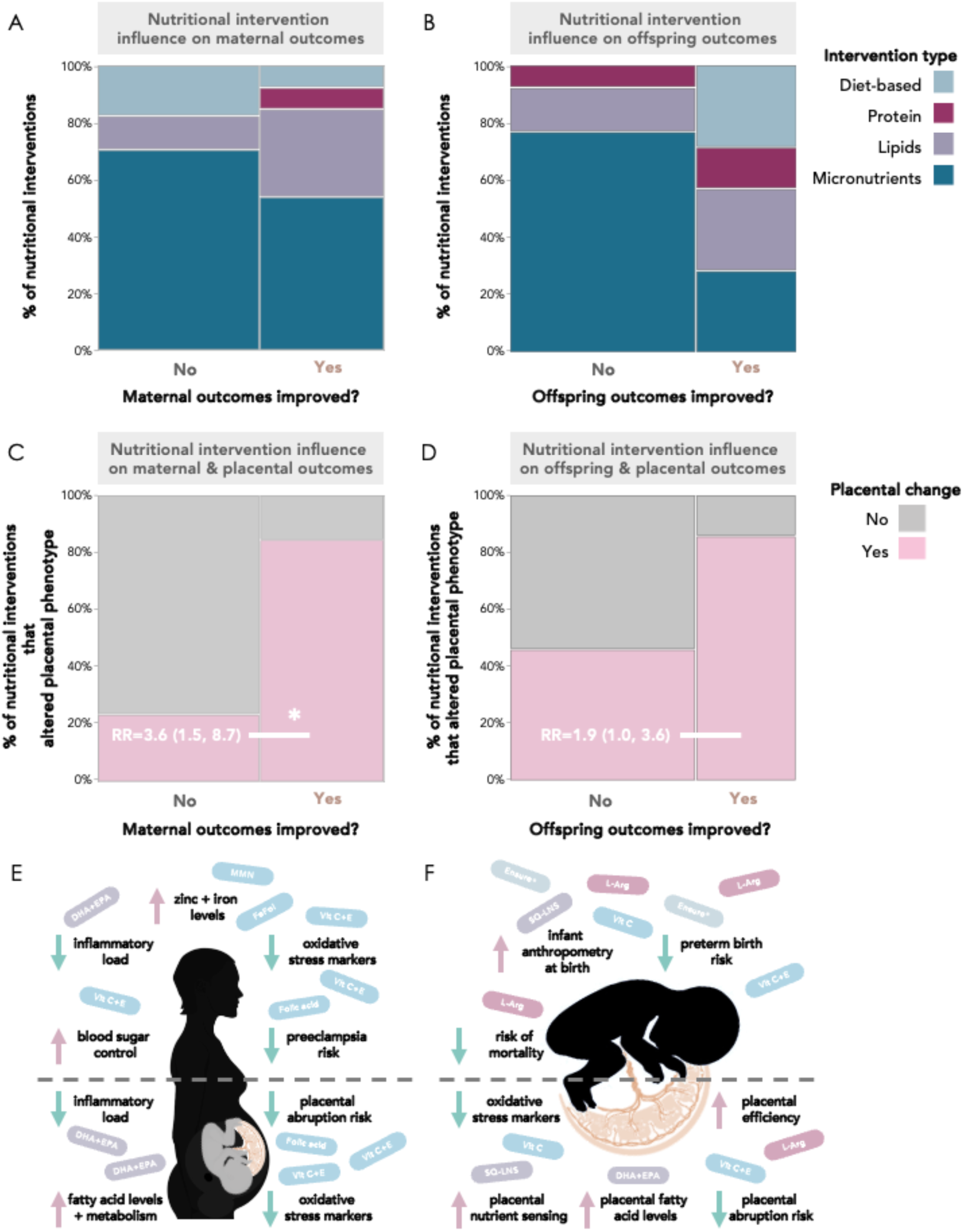
Nutritional interventions influence maternal, offspring, and placental outcomes. (A) Of the 13 nutritional interventions that associated with improved maternal outcomes, one was diet and lifestyle-based, one was protein-based, four were lipid-based, and seven were micronutrient-based. (B) Of the seven nutritional interventions that associated with improved infant outcomes, two were diet-based, one was protein-based, two were lipid-based, and two were micronutrient-based. Nutritional interventions that improved maternal (C) and offspring (D) outcomes were more likely to associate with placental changes than interventions that did not improve maternal or offspring outcomes. Visual summary of the nutritional interventions and beneficial maternal (E), offspring (F), and placental (E-F) outcomes reported in the studies under review. Mosaic plots are proportion (%) of studies reporting improvement, or no improvement, in maternal (A, C) and offspring (B, D) outcomes. Data in C and D are relative risk (95% confidence interval). *Significance = p<0.05 from Fisher’s exact test (2-tail). RR = relative risk. FeFol = iron-folic acid. Vit = vitamin. DHA = docosahexaenoic acid. EPA = eicosapentaenoic acid. SQ-LNS = small-quantity lipid-based nutrient supplements. L-Arg = L-arginine.

Results from RoB assessments were used to inform results synthesis and interpretations if the study reported significant associations between its

### Nutritional interventions associated with improved maternal and infant outcomes

Forty studies in total reported maternal and/or infant outcomes, with 50% (n=20) and 25% (n=10) of studies reporting only maternal or infant outcomes, respectively, and 25% (n=10) of studies reporting both (Supplementary Tables S5 and S6). Maternal and offspring outcomes were reported as being improved following nutritional intervention in 12/30 (40%) and 7/20 (35%) of the studies, respectively, that reported these data (Figure 5A and 5B). Descriptions of the interventions that associated with improved maternal and fetal outcomes can be found in Supplementary Results.

### Classifications of placental outcomes

Of the 53 included studies, 29 studies (51%) reported on more than one placental outcome across the five categories, for a total of 88 outcomes noted across all studies: 19 (22%) of the studies reported on placental anthropometry, 24 (27%) reported on placental molecular changes, five (6%) reported on placental pathology, 19 (22%) reported on placental abruption, and 21 (24%) reported on a placental-related disease. Key placental results from each individual study are summarized (Supplementary Table S7).

### Direct nutritional interventions associate with placental phenotype

#### Specific micronutrient-based interventions associate with placental phenotype

Of the 33 studies that used a micronutrient-based intervention, 16 (48%) reported associations between the nutritional intervention and placental phenotype (Figure 6). Micronutrient interventions that associated with altered placental phenotype began during the first (n=3), second (n=9), and third trimesters (n=4; Figure 6).

**Figure 6.**
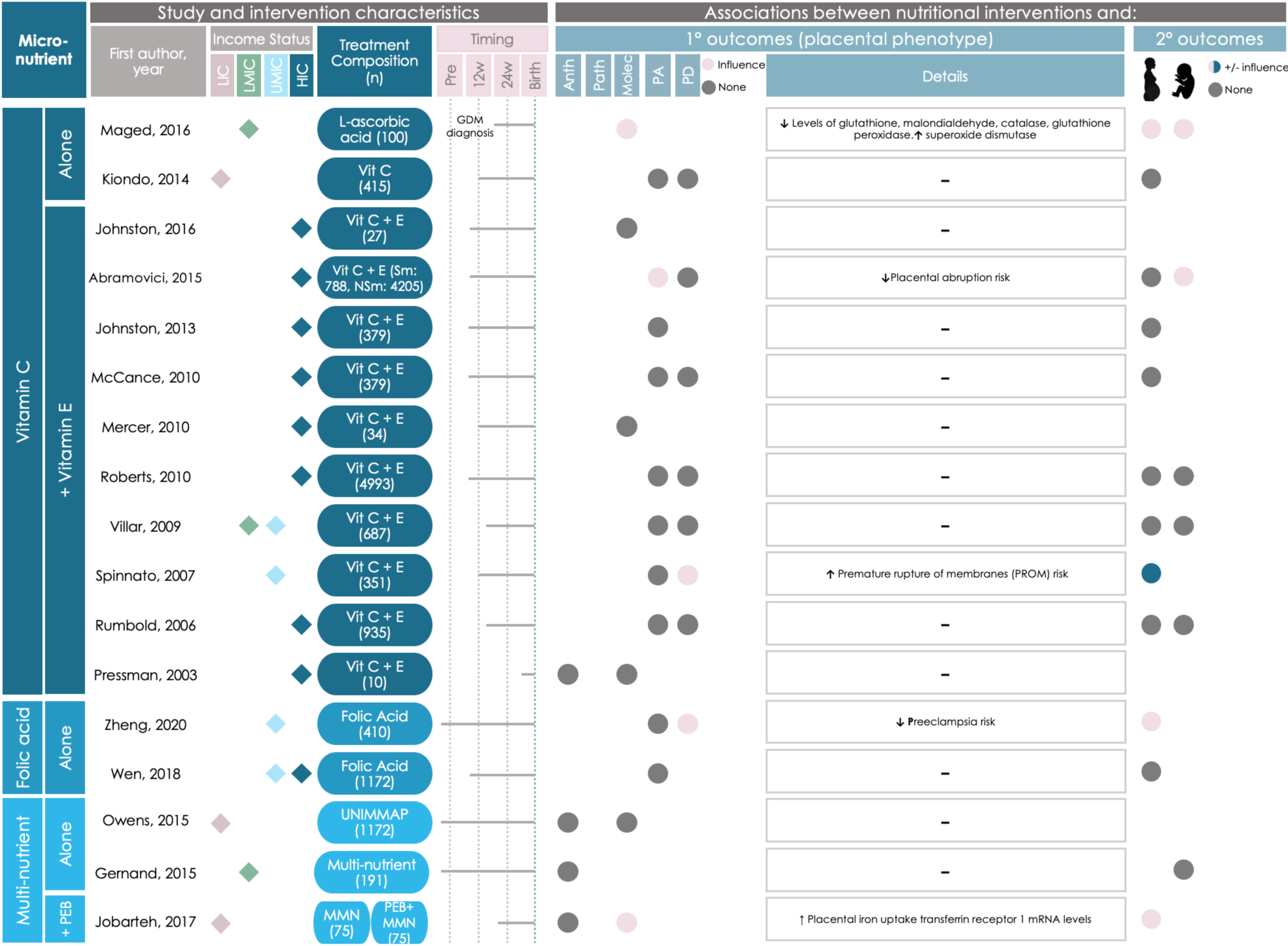
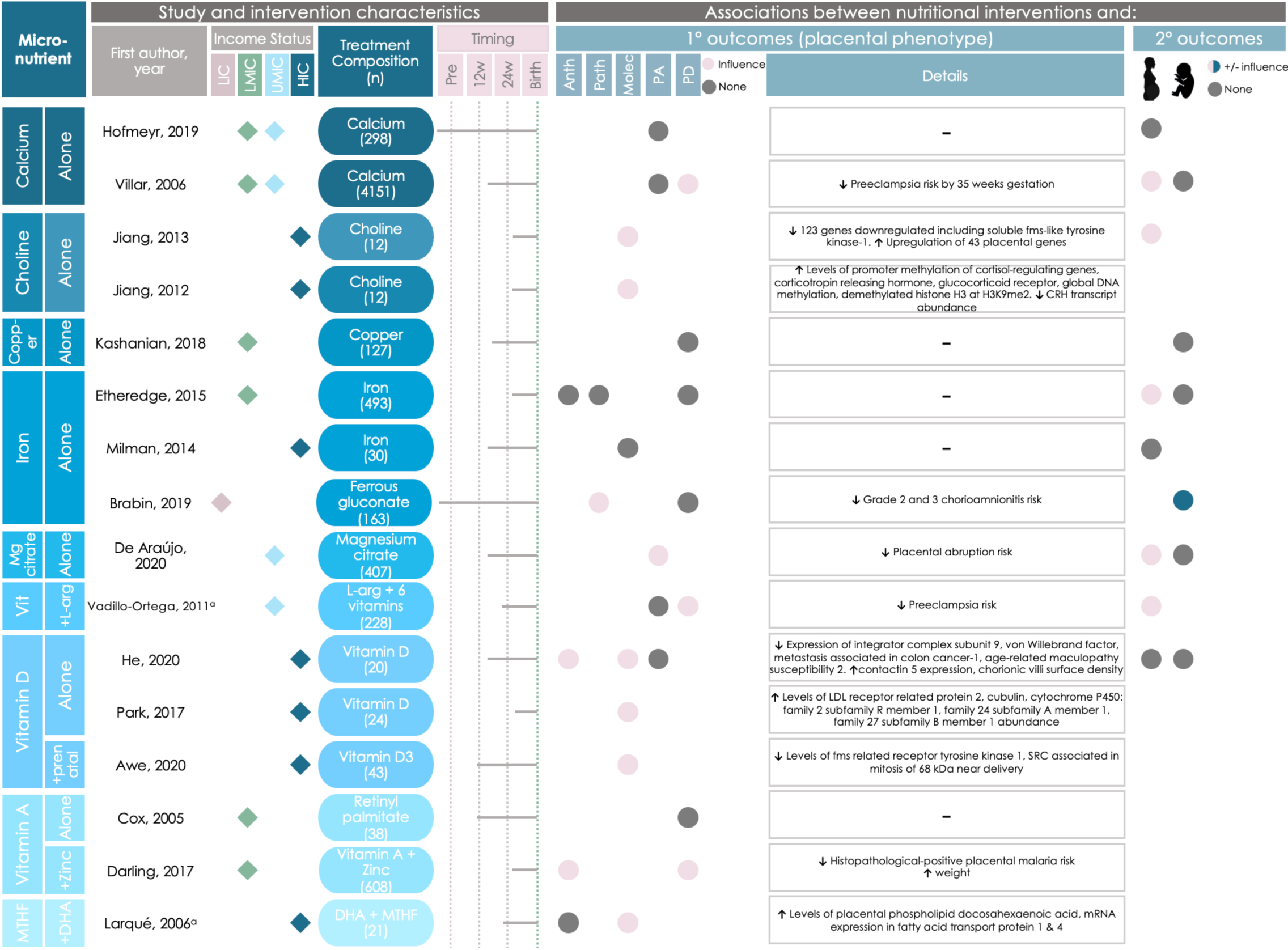
Graphical Overview for Evidence Reviews (GOfER) diagram of the impact of micronutrient-based direct maternal nutritional interventions on placental phenotypic alterations. Income status of study location obtained from World Development Indicators by The World Bank Database. LIC = Low-income country. LMIC = Low- to middle-income country, UMIC = Upper-middle-income country, HIC = High-income country. Sm = Smoker. NSm = Non-smoker. N = Number of participants. Pre = Pre-conception. PEB = Protein-Energy Ball. MMN = Multiple Micronutrient. GDM = Gestational diabetes mellitus. Anth = Anthropometry. Molec = Molecular Alterations. Patho = Pathology. PA = Placental abruption. PD = Placenta-related disease. A dash (-) under “Details” indicates no significant placental phenotype alterations. Circles for this review’s secondary outcomes were only placed if the study reported fetal/infant or maternal outcomes as a primary outcome of their study. ^a^Vadillo-Ortega, 2011, and Larqué, 2006 appear as duplicates on 2 separate pages between figures 6-8 due to their crossover between intervention categories. Income status of study location obtained from World Development Indicators by The World Bank Database. LIC = Low-income country. LMIC = Low- to middle-income country, UMIC = Upper-middle-income country, HIC = High-income country. N = Number of participants. Pre = Pre-conception. L-arg = L-arginine. MTHF = 5-methyl-tetrahydrofolic acid. Anth = Anthropometry. Molec = Molecular Alterations. Patho = Pathology. PA = Placental abruption. PD = Placenta-related disease. A dash (-) under “Details” indicates no significant placental phenotype alterations. Circles for this review’s secondary outcomes were only placed if the study reported fetal/infant or maternal outcomes as a primary outcome of their study. ^a^Vadillo-Ortega, 2011, and Larqué, 2006 appear as duplicates on 2 separate pages between figures 6-8 due to their crossover between intervention categories.

Associations between micronutrient-based nutritional interventions and placental anthropometry were reported in two studies, including increased placental weight at term (following daily vitamin C and zinc supplementation initiated during the first trimester in a population at high risk for malaria and low birthweight (32)), and increased chorionic villi density (following vitamin D supplementation that began in the late first or early second trimester in mothers at high risk of having offspring with asthma (33); Figure 6).

One study reported associations between micronutrient-based interventions and placental pathology (34), where daily iron supplementation initiated periconceptionally was associated with a decreased risk of chorioamnionitis (a maternal inflammatory response leading to the infiltration of neutrophils into the placenta) in a population at high risk for malaria and chorioamnionitis (34) (Figure 6).

Half (n=8) of these 16 micronutrient-based intervention studies reported differences in placental molecular changes between the control and intervention groups. Placental molecular changes included increased expression or activity of the following: placental superoxide dismutase (an antioxidant enzyme (35); following daily vitamin C supplementation initiated upon gestational diabetes diagnosis (36)), transferrin receptor 1 mRNA (involved in placental iron uptake (37, 38); following a daily multiple micronutrient intervention that began in the second trimester (38)), genes involved in vitamin D metabolism (after daily vitamin D supplementation that started in the third trimester (39)), soluble fms-like tyrosine kinase-1 (SFLT-1; following daily vitamin D plus 5-methyl-tetrahydrofolic acid [MTHF] supplementation that started in the second semester(40)), and placental phospholipid DHA (after daily 5-MTHF and DHA supplementation that began in the second trimester (41)). Molecular changes also included modified expression of five genes in placental tissue (increased expression of metastasis associated in colon cancer-1 [*MACC1*], integrator complex subunit 9 [*INST9*], Von Willebrand factor [*vWF*], and age-related maculopathy susceptibility 2 [*ARMS2*]; and decreased expression of contactin 5 [*CNTN5*]; following daily vitamin D supplementation initiated in the second trimester (33)). In two studies from a single trial of daily maternal choline supplementation initiated during the third trimester (42), increased methylation of multiple cortisol-regulating genes and dysregulation of 197 different placental biological processes was reported (43, 44).

Placental abruption risk following nutritional intervention was assessed in 16 studies, where two interventions, which supplemented either daily vitamin C+E (45) (initiated during the first trimester) or magnesium citrate (46) (initiated during the second trimester) were associated with decreased abruption risk (Figure 6). In contrast, two studies from the Diabetes and Preeclampsia Intervention Trial (DAPIT) (47), which supplemented daily vitamin C+E (that began in the first trimester) in mothers with diabetes to lessen preeclampsia risk, reported no differences in placental abruption risk in the full cohort (47) or in follow-up studies with a cohort subset (48). Two studies from the Combined Antioxidant and Preeclampsia Prediction Studies (CAPPS) trial reported that placental abruption risk was decreased in mothers who smoked (45), but not the intervention group overall (49), following daily vitamin C+E supplementation that started in the first trimester.

Placental-related disease risk was altered in 5 (33%) of the 15 studies that evaluated placental-related disease occurrence following nutritional intervention. These findings included increased risk of premature rupture of membranes (PROM; following daily vitamin C+E supplementation initiated in the second trimester (50)), and a decreased risk of histopathological-positive placental malaria risk (following daily vitamin A and zinc supplementation that started in the third trimester (32)), and preeclampsia (following daily interventions with either folic acid (51) that began pre-conceptionally, or calcium (52) initiated in the second trimester; Figure 6). In the CAPPS trial (53), no changes in the risk of preeclampsia (45, 49), or amnio-choriodecidua (54), were reported following daily vitamin C+E supplementation initiated in the first trimester.

For the remaining 17/33 studies that used a micronutrient-based intervention, there were no reported associations between micronutrient interventions and placental anthropometry (n=5 [30%]) (55–58), pathology (n=1 [6%]) (58), molecular changes (n=5 [30%]) (54–56, 59, 60), abruption risk (n=8 [47%]) (47–49, 61–65), or placental-related disease (n=8 [47%]) (47, 49, 58, 61–63, 66, 67).

#### Specific macronutrient-based interventions influence placental phenotype

Of the 11 studies that reported on lipid-based interventions, nine (82%) reported placental changes in the intervention group (Figure 7). Lipid-based interventions that associated with altered placental phenotype began during the first (n=2), second (n=5), and third trimesters (n=2; Figure 7).

**Figure 7.**
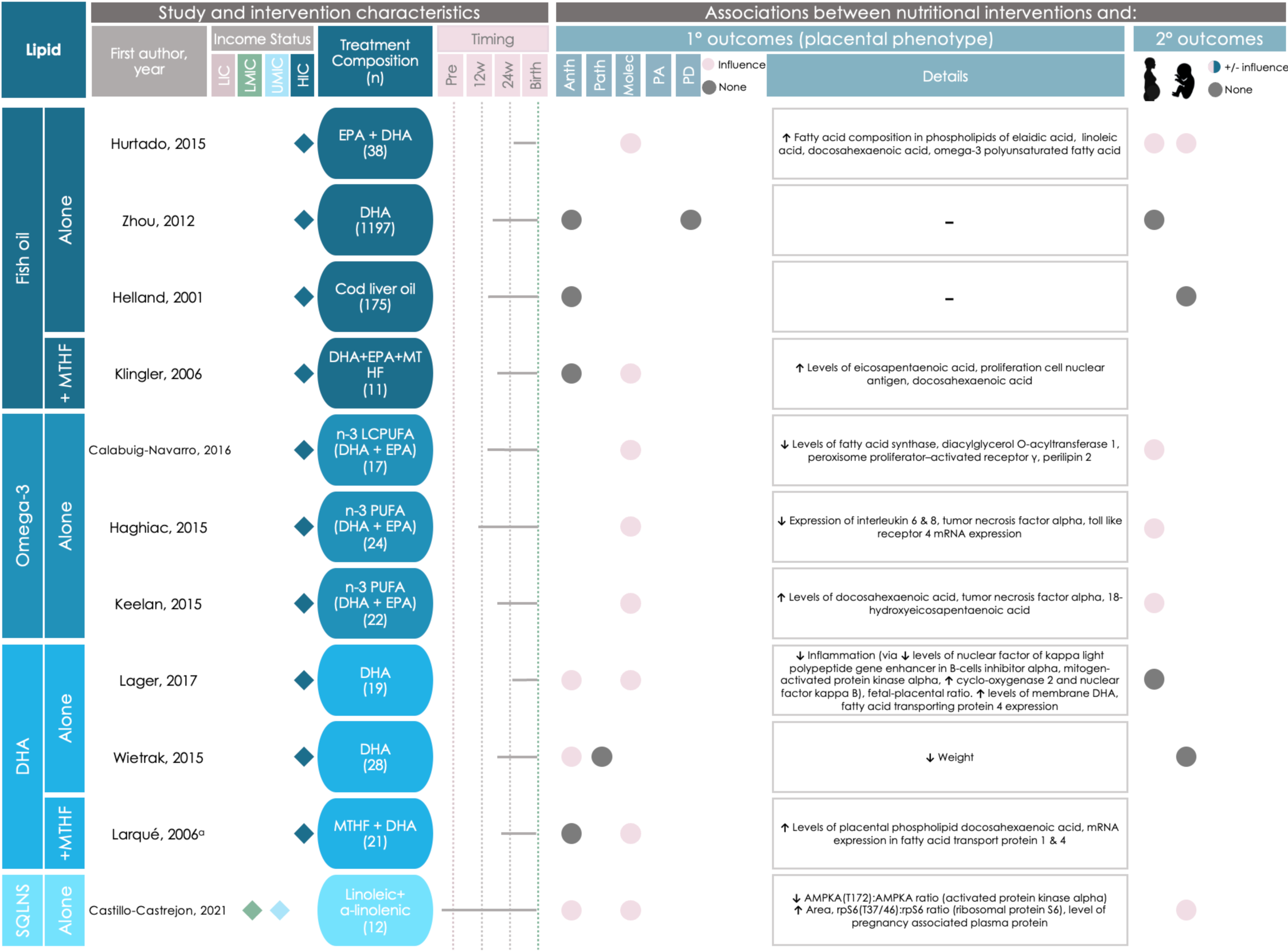
Graphical Overview for Evidence Reviews (GOfER) diagram of the impact of lipid-based direct maternal nutritional interventions on placental phenotypic alterations. Income status of study location obtained from World Development Indicators by The World Bank Database. LIC = Low-income country. LMIC = Low- to middle-income country, UMIC = Upper-middle-income country, HIC = High-income country. N = Number of participants. Pre = Pre-conception. L-arg = L-arginine. MTHF = 5-methyl-tetrahydrofolic acid. SQLNS = Preconceptional maternal small-quantity lipid-based nutrient supplementation. EPA = eicosapentaenoic acid. DHA = docosahexaenoic acid. N-3 PUFA = omega-3 polyunsaturated fatty acid. Anth = Anthropometry. Molec = Molecular Alterations. Patho = Pathology. PA = Placental abruption. PD = Placenta-related disease. A dash (-) under “Details” indicates no significant placental phenotype alterations. Circles for this review’s secondary outcomes were only placed if the study reported fetal/infant or maternal outcomes as a primary outcome of their study. ^a^Vadillo-Ortega, 2011, and Larqué, 2006 appear as duplicates on 2 separate pages between figures 6-8 due to their crossover between intervention categories.

Changes in placental anthropometry were reported in three lipid-based intervention studies (Figure 7). Decreased placental weight (68) and fetal:placental weight ratio (69) were reported following daily DHA supplementation that began in the second and third trimester, respectively, while pre-conceptionally initiated daily maternal small-quantity lipid-based nutrient supplementation (SQLNS) was associated with an increase in placental area (70) (Figure 7).

Eight lipid-based interventions were associated with molecular changes in the placenta, namely in placental processes related to inflammation and fatty acid transport (Figure 7). The molecular changes included increased expression or concentration of: placental linoleic acid (following daily fish oil supplementation that started in the third trimester (71)), proliferating cell nuclear antigen (PCNA; which plays a key role in nucleic acid metabolism (72); following daily fish oil supplementation that began in the second trimester (73)), and tumour necrosis factor alpha (*TNFα*; following daily omega-3 supplementation that started in the second trimester (74)). Decreased expression of placental inflammatory markers (following daily DHA supplementation initiated in the third trimester (69)), and decreased activated protein kinase alpha ratio (AMPKA [a regulator in cell metabolism]; following daily pre-conceptionally initiated SQLNS supplementation (70)) were also reported (Figure 7). Two studies from a single clinical trial that supplemented omega-3 fatty acids (beginning in the second trimester) reported a significant reduction in the expression of pro-inflammatory genes in placental tissue and decreased placental lipid storage capacity (75–77). Two studies that reported findings from a trial of daily DHA and MTHF supplementation that began in the second trimester reported alterations to DHA content in placental phospholipids and expression of PCNA (73) and alterations to placental fatty acid transport proteins (41).

Two of the 11 (18%) lipid-based interventions did not associate with placental changes (Figure 7). In both interventions, a daily fish oil supplement was taken, beginning in the second trimester, and placental weight (78) and preeclampsia risk (79) were not altered (Figure 7).

Three of the 53 studies used amino acid- or protein-based interventions (Figure 8). Two (66%) of these three studies reported associations between the intervention and placental phenotype. Daily supplementation with L-arginine initiated during the third trimester was associated with an increased cerebro-placental ratio (a gestational indicator associated with adverse pregnancy outcomes like low-birth weight (80)) when compared to the control group (81), and twice daily supplementation with L-arginine and antioxidant vitamins that began in the second trimester associated with a decreased risk of preeclampsia (82) (Figure 8).

**Figure 8.**
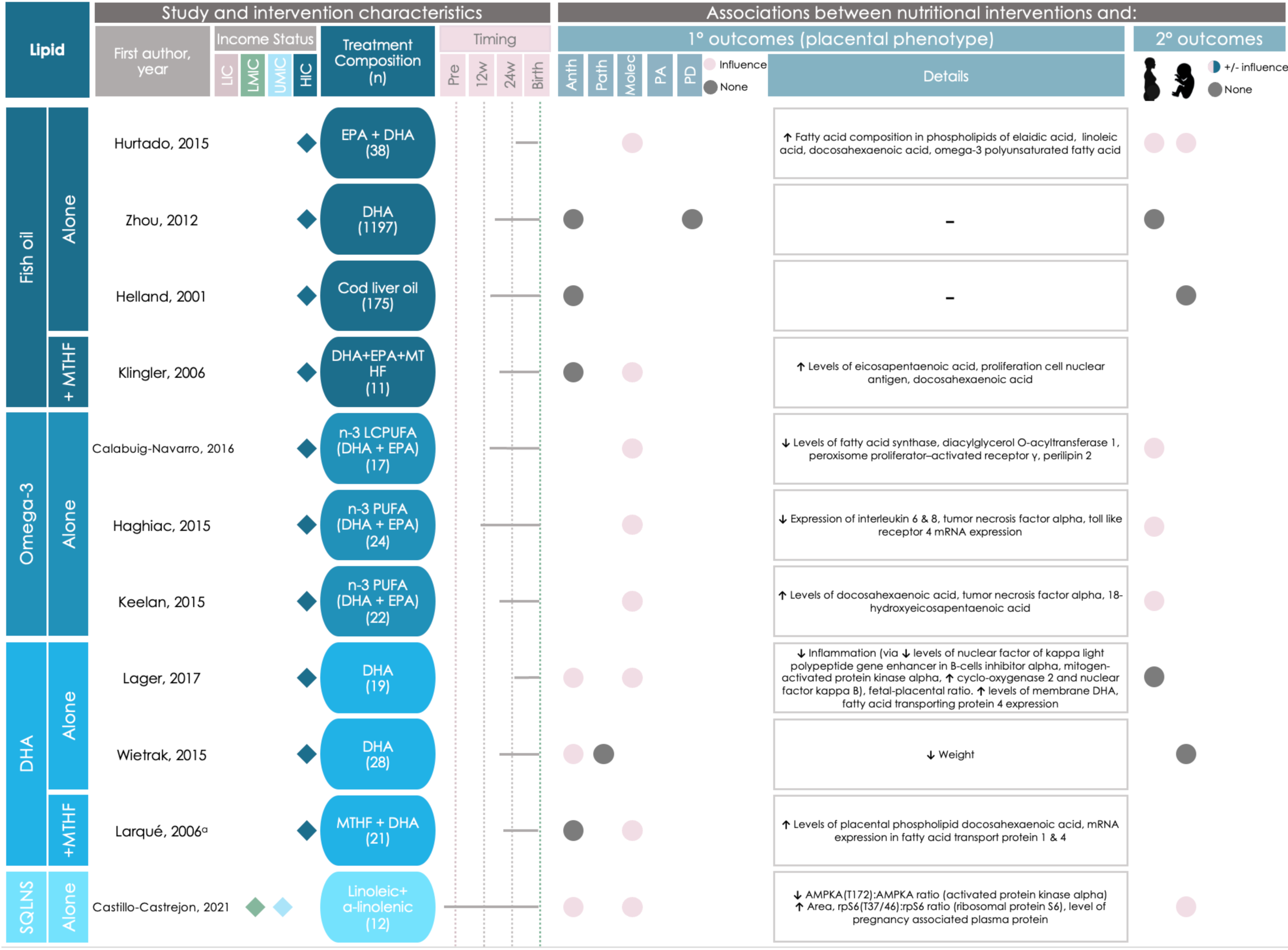
Graphical Overview for Evidence Reviews (GOfER) diagram of the impact of protein- and diet and lifestyle-based direct maternal nutritional interventions on placental phenotypic alterations. Income status of study location obtained from World Development Indicators by The World Bank Database. LIC = Low-income country. LMIC = Low- to middle-income country, UMIC = Upper-middle-income country, HIC = High-income country. N = Number of participants. Pre = Pre-conception. L-arg = L-arginine. IPTp-SP = intermittent preventative treatment in pregnancy with sulphadoxine-pyrimethamine. DiMO = monochorionic diamniotic twin pregnancy. Anth = Anthropometry. Molec = Molecular Alterations. Patho = Pathology. PA = Placental abruption. PD = Placenta-related disease. A dash (-) under “Details” indicates no significant placental phenotype alterations. Circles for this review’s secondary outcomes were only placed if the study reported fetal/infant or maternal outcomes as a primary outcome of their study. ^a^Vadillo-Ortega, 2011, and Larqué, 2006 appear as duplicates on 2 separate pages between figures 6-8 due to their crossover between intervention categories.

The one amino acid- or protein-based intervention that did not associate with placental phenotype was supplementation of L-arginine initiated in the third trimester, which did not associate with changes in placental weight, abruption risk, or thrombosis (83) (Figure 8).

#### Specific diet and lifestyle-based interventions influence placental phenotype

Eight interventions were diet and lifestyle-based and three (38%) associated with placental phenotype.

An intervention that delivered a daily probiotic supplement and dietary education and support (to promote a diet in line with nutritional guidelines and maintenance of appropriate fat consumption) that began during the second trimester associated with increased concentration of polyunsaturated fatty acids in the placenta (84); Figure 8).

Two diet and lifestyle-based interventions were associated with a reduced risk of placental-disease. Daily Ensure^®^ liquid nutritional supplementation initiated during the second trimester was associated with reduced TTTS diagnosis and incidence at birth (31) (Figure 8). Haematinics and intermittent preventative treatment (IPTp-SP) administered daily during the third trimester until birth was associated with reduced placental malaria incidence in the treatment group when comparing mothers with the same human immunodeficiency virus (HIV) status (85) (Figure 8). Notably, both of these studies had a moderate RoB due to potential deviations from the intended interventions (31, 85) and one had a serious RoB due to the methods used for selection of participants into the study (31).

The remaining five (63%) of diet and lifestyle-based interventions did not associate with placental changes (Figure 8) (30, 86–89).

### Intervention effects on the placenta vary by study location

Studies that took place in LIC and LMIC reported fewer associations between nutritional interventions and placental changes than studies that took place in UMIC and HIC (Figure 3A). All nutritional interventions in LIC (n=4) were micronutrient-based (Figure 3B). Of these, two (50%) reported associations between the intervention and placental pathology (n=1 study) and placental molecular phenotype (n=1; Figure 3A). Nutritional interventions in LMIC were micronutrient-based (n=9 [75%]), lipid-based (n=1 [8%]), or diet and lifestyle-based (n=2 [16%]), of which, five (42%) reported placental changes following the nutritional intervention (Figures 3A-B). In the five LMIC-based studies, associations between the nutritional intervention and placental anthropometry (in n=2 studies [29%]), placental molecular phenotype (n=2 [29%]), and placenta-related disease (n=3 [43%]) were reported.

Nutritional interventions in UMIC were micronutrient-based (n=8 [89%]) or lipid-based (n=1 [11%]), and of these, six (66%) reported associations between the intervention and placental phenotype (Figures 3A-B). In these six UMIC studies, associations between the nutritional intervention and placental anthropometry (in n=1 study [13%]), placental molecular phenotype (n=1 [13%]), placental abruption (n=1 [13%]), and placental-related disease (n=5 [63%]) were reported.

Nutritional interventions in HIC were micronutrient-based (n=16 [48%]), lipid-based (n=16 [48%]), protein-based (n=2 [6%]), or diet and lifestyle-based (n=6 [18%], Figure 3B), of which, 51% (n=17) reported associations between the intervention and placental phenotype (Figure 3A). These 17 HIC-based studies reported associations between the nutritional intervention and placental anthropometry (in n=4 studies [20%]), placental molecular phenotype (n=14 [70%]), placental abruption (n=1 [5%]), and placental-related disease (n=1 [5%]).

### Interventions associated with placental phenotype are more likely, overall, to improve maternal and offspring outcomes

Maternal outcomes were more likely to be improved in studies that reported placental changes following nutritional intervention than studies that reported no placental changes (n=11 [73%] vs. n=2 [13%], RR = 3.6 [1.5, 8.7]; Figure 5C).

Studies that reported placental changes were more likely to report improved offspring outcomes than those that did not report placental changes (n=6 vs. n=1, RR = 1.9 [1.0, 3.6]; Figure 5D).

More details on the nutritional interventions that improved maternal and offspring outcomes are provided in the Supplementary Results.

### Data on sex differences in placental response to nutritional interventions are lacking

Only 17 (32%) of the studies under review included data on the number of male and female offspring in the study, and no studies reported data on placental outcomes stratified by sex.

## Discussion

Here, we reviewed evidence on how direct maternal nutritional interventions influence placental phenotype in humans. Lipid-based interventions were most likely to associate with altered placental phenotype, often related to changes in inflammatory processes or markers. We also found that interventions associated with placental changes were more likely to also improve maternal and offspring outcomes. Studies conducted in HICs were the most likely to report associations between nutritional interventions and placental phenotype, although LICs were underrepresented in the reviewed studies. There was a lack of reporting on sex-differences or -specific effects of nutritional interventions on placental outcomes. Investigating how direct maternal nutritional interventions during preconception and pregnancy influence placental phenotype strengthens our understanding of the mechanisms underlying intervention effectiveness and could lead to more targeted approaches to improve maternal-fetal health.

Nutritional interventions associated with placental changes were more likely to also improve maternal and offspring outcomes. Specifically, improved maternal iron levels, inflammatory status, and blood sugar control may be accompanied by placental changes such as increased nutrient transporter expression (38) and decreased inflammation and oxidative stress (36, 75). Similarly, decreased placental abruption risk (45), improved cerebro-placental ratio (81), and greater placental area (70) following nutritional intervention corresponded with a decreased risk of preterm birth and IUGR and improved fetal growth. While it is well-established that nutritional interventions can improve maternal and offspring outcomes (90, 91), our findings suggest that understanding the placental response to nutritional interventions may reveal insights into the biological mechanisms underlying their effects on maternal or offspring health.

Several studies reported changes in maternal, placental, or offspring levels of pro-inflammatory biomarkers following micronutrient- and lipid-based nutritional interventions (36, 41, 69, 71, 73–75). For example, supplementation with vitamin C, which has antioxidative properties (92), associated with reduced levels of oxidative stress markers in placental tissue and maternal and offspring blood (36). Excess oxidative stress in the placenta can promote inflammation and adversely affect fetoplacental development and future disease risk (93–96). Lipid-based interventions associated with increased anti-inflammatory DHA and EPA levels in the placenta and maternal circulation (41, 69, 71, 73, 74). During pregnancy, DHA and EPA promote angiogenesis and reduce oxidative stress and inflammation in the placenta, and are required for fetal brain development (97). These findings collectively suggest that common nutritional interventions can impact placental inflammatory processes, which may in turn influence maternal and offspring outcomes (94–96). Notably, two of these studies had a high RoB due to blinding of participants or study personnel (36, 69), while two others had an unclear RoB in half of the assessment categories due to inadequate information in the publications (41, 73). High quality studies, that adhere to best practice reporting standards, are needed to better understand how maternal nutritional interventions affect placental inflammation, especially in pregnancies with maternal comorbidities like obesity and metabolic syndrome, which are associated with chronic inflammation and altered placental nutrient transport (98, 99).

None of the reviewed studies reported placental changes by sex. Given the sex-specific differences in placental development, including in nutrient exchange capacities (100), it is likely that placental sex influences the placental response to nutritional interventions (101) and this is an area requiring further research. Additionally, LICs were underrepresented in the reviewed studies, with only four studies taking place in LICs. As LICs often face higher rates of undernutrition and are where nutritional interventions are most needed (102, 103), further research on how nutritional interventions impact the placenta in these regions is needed.

Strengths of this study include it being the first to synthesise knowledge on the relationships between direct maternal nutritional interventions and placental phenotype, extending beyond placental-related pregnancy complications (104). Further, our focus on only human studies increases the clinical relevancy of our findings (105), and we identified key gaps in the literature, such as the lack of sex-stratified placental data and underrepresentation of LICs and LMICs, which may guide future research. The limited placental data from nutritional interventions in low-resource settings reflects challenges in implementing such interventions in these contexts more broadly, which often stem from issues related to resource availability and mobilization (106). A lack of standardised placental biobanking is another barrier to studying the placental impacts of nutritional interventions in low-resource settings. Ongoing biobanking initiatives, such as the Global Pregnancy Collaboration (CoLab), the PREgnancy Care Integrating Translational Science Everywhere (PRECISE) Network, and the Alliance for Maternal and Newborn Health Improvement (AMANHI) study (107–109), aim to address this gap.

Limitations of this review first include the lack of formal meta-analysis due to the high variability in the types of placental outcomes reported in the reviewed studies. In many cases, the scope and detail of placental data available in the reviewed studies were limited and there was high heterogeneity in how placental outcomes were reported. Second, we were unable to assess how individual-level factors like dietary history, nutritional status, or fetoplacental sex influenced the placental response to an intervention, as these data were not available in the reviewed studies. The lack of measurement or adjustment for these key factors is therefore a likely source of bias across the reviewed studies. Third, intervention type, timing, frequency, and duration also varied, which prevented direct comparisons between studies. Fourth, the reviewed studies were largely concentrated in HICs, limiting the generalizability of our findings to other contexts, and fifth, around one quarter of studies did not report or had unclear compliance data, which may have influenced the intervention’s impact on placental phenotype. Lastly, while we have conservatively highlighted consistent themes from the reviewed studies, it’s important to recognize that results varied widely even among studies with relatively similar interventions and outcomes. With the available evidence, it is therefore not currently possible to draw conclusions on the strength of associations observed. If future studies collect and report standardised outcomes and the evidence base on how nutrition interventions influence the placenta expands, future reviews can adopt a more focused approach to better understand the mechanisms that may contribute to success of specific intervention types and their effects on the placenta.

### Advancing nutritional interventions and maternal-child health through placental research

Informed by the findings of this review and elaborating on recommendations from other groups (107–109), we propose the expansion of the existing core outcome set, “*Pregnancy Nutrition Core Outcome Set”* (110), to include the following new variables:

1. Fetal/placental sex
2. Placental anthropometry (weight, diameter)
3. Placental pathology (where applicable, or banking of fixed placental specimens for future histomorphology and pathology assessments)
4. Adverse events and effects related to the placenta
5. Collection of placental tissue core samples for future analysis of placental phenotype, including function, if not intended for the immediate study

Standardised reporting of placental and maternal variables is essential for understanding placental responses to nutritional interventions, improving comparability across trials, and reducing outcome heterogeneity. These factors limited our ability to make direct comparisons in this review and have been recognized by others as challenges that core outcome sets can help address (111).

We also recommend increased collection and phenotyping of placental samples from nutritional intervention studies in LMICs and LICs. Aligning with efforts to promote standardized placental biobanking like CoLab, the PRECISE Network, and the AMANHI study (107–109), this would improve representation and understanding of placental mechanisms and adaptations to nutritional exposures in diverse populations and geographies.

Finally, future research should focus on determining optimal windows for nutritional interventions (i.e., interventions initiated pre-conceptionally, in early pregnancy, or in later pregnancy) for improving maternal and offspring outcomes, and how the placenta adapts to, and influences pregnancy outcomes, based on timing of exposure. Research focused on developing human nutritional intervention(s) targeted to the placenta to, in turn, improve pregnancy/offspring outcomes, should also be a priority. Non-nutritional based therapies (such as the use of liposomes to enhance drug delivery to the placenta) have the potential to improve offspring outcomes via placental changes (112), and nutritional interventions may be able to do the same.

### Conclusions

We found that direct maternal preconception and prenatal nutritional interventions that target maternal health and nutritional status also influence placental development and function. These (potentially unintended) placental adaptations are likely beneficial for the pregnancy and may underlie the improved growth and development of offspring observed in these pregnancies. Understanding how the placenta responds to nutritional interventions is key for modifying and improving strategies to address nutrient deficiencies, reducing maternal and neonatal morbidity and mortality and improving offspring health trajectories globally.

## Supporting information

Supplementary Results

## Data Availability

All data discussed are available in the published articles included in this review.

## Author contributions

Conceptualization, MW and KLC.; methodology, VB, MW, and KLC; formal analysis, VB and MW; investigation, VB, MW, and KLC; data curation, VB and MW; writing—original draft preparation, VB, MW, and KLC; writing—review and editing, VB, MW, and KLC; visualisation, VB, MW, and KLC; supervision, KLC and MW; funding, KLC. All authors have read and agreed to the published version of the manuscript.

