## Supplementary Methods for "Do nutritional interventions before or during pregnancy affect placental phenotype? Findings from a systematic review of human clinical trials"

*Information sources and search terms*

PubMed, ClinicalTrials.gov, and the World Health Organization (WHO) International Clinical Trials Registry Platform (ICTRP) were searched to identify peer-reviewed publications using the following search string: ((nutr* OR diet* OR supplement* OR vitamin OR folate* OR folic acid OR mineral OR (micronutri* AND (intervention OR supplement*))) AND (placenta* OR preeclampsia) AND (maternal OR pregnan* OR gravid* OR conception* OR preconceptional OR gestation*)). When searching the listed databases, the filters "Clinical Study, Clinical Trial, Comparative Study, Controlled Clinical Trial, Journal Article, Humans" were applied.

*Article search, screening, and data collection*

A three-level screening process was performed by two authors (VB and MW; Figure 1). A total of 5299 titles from PubMed (n=5078), ClinicalTrials.gov (n=156), and WHO ICTRP (n=65) were captured from the article search. Articles were excluded for deduplication (n=14), publication date before 2001 (n=8), and incorrect study type (n=10), leaving 5267 articles to be screened at level one (title screening). Clinical trial titles from ClinicalTrials.gov and WHO ICTRP, and article titles from PubMed search results were screened for relevance to our review objectives. In total, 4568 titles were excluded for having a topic not relevant to this review, leaving a total of 699 article abstracts and clinical trials documents to be screened at level two.

At screening level two, 699 article abstracts (PubMed: n=608) and clinical trial records (ClinicalTrials.gov: n=60, WHO ICTRP: n=31) were reviewed to determine their adherence to the inclusion criteria (Figure 1). Articles and trials were excluded for: not reporting on a direct maternal nutritional intervention (n=323), reporting on an animal study (n=12), incorrect type of article (i.e., review, study protocol, or commentary; n=169), no results posted (n=29), or not written in English (n=2), leaving 164 records (PubMed articles: n=129, ClinicalTrials.gov: n=17, WHO ICTRP: n=18) to be carried forward to level three screening.

At level three, full texts were unavailable for seven of 129 PubMed articles. The corresponding authors were contacted by email twice between December 2021 and January 2022 for each of these seven articles, but only one response was received. Publications resulting from the clinical trials retained for screening at level three were obtained from the trial webpage directly, or from searching the trial name and/or number using search browsers (e.g., Google Scholar). Ultimately, 158 full texts were reviewed to evaluate whether studies met the full inclusion criteria (from PubMed, n=123; ClinicalTrials.gov, n=17, and WHO ICTRP, n=18; Figure 1). At level three, articles were excluded for not including a placental measure (n=64; including articles reporting on relationships between a nutritional intervention and risk of preeclampsia that did not provide placenta-specific measures), not administering a direct nutritional intervention (n=9), duplicates (n=11), or other reasons (i.e., wrong study type, no trial results available or could not be found (n=21). In total, 53 articles met the full inclusion criteria.

*Data extraction*

Data were extracted from each of the 53 articles, including population data (such as study location [which we used to identify socioeconomic status based on four World Bank classifications: low-income (LIC), low-middle income (LMIC), upper-middle income (UMIC), and high-income

(HIC)] (1)), maternal clinical characteristics and demographics, type of pregnancy (singleton or twin), and nutritional intervention details (type [micronutrient, lipid, protein, and/or diet- or lifestyle-based], composition, timing, dose, and compliance data). Data on maternal comorbidities (defined in this study as conditions or states of health during pregnancy that may be harmful to the health of the mother and/or fetus) reported in the studies under review were noted and used to inform results interpretations. Key results from each individual study were extracted and summarized, and the proportion of studies that did or did not report placental changes or improvements in maternal and offspring outcomes was calculated for each subtype of nutritional intervention (micronutrient, macronutrient, and diet and lifestyle-based). Reported adverse outcomes were noted and classified into two categories: adverse effects (outcomes that were suspected to be in response to the intervention (2)) and adverse events (outcomes that were not suspected to be in response to the intervention (2)). Data on associations between the nutritional intervention and 1. placental phenotype (primary outcome), and 2. placental sex, and 3. fetal/infant and maternal outcomes (secondary outcomes) were also captured.

*Risk of bias assessments*

The articles under review were assessed for risk of bias (RoB) using the Cochrane Collaboration's Tool for Assessing Risk of Bias for randomised studies (n=50; Supplementary Table S3) and the Risk Of Bias In Non-randomised Studies of Interventions (ROBINS-I) tool for non-randomised studies (n=3) (3, 4) .The RoB assessment criteria were set *a priori*. As our outcome of interest was placental phenotype, which was often not the primary outcome of the studies under review, assessments for risk of bias due to "incomplete outcome data" considered (when applicable) how the subset of the original cohort with placental data was selected or determined. We also assessed potential bias related to compliance to the intervention, as follows: articles that clearly measured and reported participant compliance were assessed as low risk, articles that did not report clear methods to measure compliance, but had frequent follow up visits with participants, were assessed as unclear risk, and articles that did not measure compliance and had few participant follow up visits (e.g., only at intervention onset and delivery) were assessed as high risk. RoB assessments were performed independently by two authors (VB and MW). Discrepancies in RoB assessments were resolved through discussion with a third author (KLC). While RoB in interventions was not a primary outcome of this review, assessments of RoB were performed to inform results interpretations.

*Data synthesis and visualisation*

Graphical Overview for Evidence Reviews (GOfER (5)) figures were created for each type of nutritional intervention to visualise key study data. The GOfER included intervention composition and timing, reported alterations to placental phenotype, and reported maternal and fetal outcomes (both beneficial and adverse). A filled map was created to demonstrate the locations of the studies included (Microsoft Excel v16). Alluvial diagrams were created to visualise relationships between type of intervention, study location, and changes in placental phenotype (or no changes; RAWGraphs (6)).
