## Supplementary Results for "Do nutritional interventions before or during pregnancy affect placental phenotype? Findings from a systematic review of human clinical trials"

*Study and cohort characteristics*

The majority (n=46 [87%]) of studies included only singleton pregnancies, while seven (13%) included both singleton and twin pregnancies. Maternal age in the included studies ranged from 15 to 45 years old, with most studies reporting an average maternal age in the mid- to late twenties. Five studies enrolled mothers under the age of 18 (1-5). Maternal comorbidities were common in the studies under review (n=33 [62%]) and were either a criterion to be a participant in the nutritional intervention trial or were identified as a potential confounding variable by the original authors. The most common comorbidities included a history or diagnosis of preeclampsia (n=12 [36%]), prior or current hypertension (n=7 [21%]), overweight and/or obesity (n=7 [21%]), smoking (n=6 [18%]), and/or any type of diabetes (n=6 [18%]; Supplementary Table S4). Twenty studies excluded pregnant participants with comorbidities.

Presence of a maternal nutrient deficiency was a selection criterion for three (5.6%) of the studies. All studies reported self-administration of the intervention by the participant, and three studies also included an additional intervention administered by a research team member once weekly. Interventions began either peri- (n=3 [6%]) or post-conceptionally (n=50 [94%]; 14 beginning in the first trimester, 28 in the second trimester, and 11 in the third trimester), with the majority continuing until birth (n=52 [98%]), and one that finished after 12 weeks of administration.

Adverse effects and events were reported in five and one study, respectively, and included gastrointestinal complaints following multiple micronutrient (3) or iron (6) supplementation, an increased incidence of preterm birth following iron supplementation (1), an increased likelihood of antenatal hospital admission with hypertension following vitamin C+E supplementation (exact cause unknown to the researchers) (7), and increased PROM risk and reports of general developmental abnormalities in both the treatment vitamin C+E and placebo groups (8).

*Nutritional interventions associated with improved maternal and infant outcomes*

There were 13 nutritional interventions that associated with improved maternal outcomes, including one diet and lifestyle-based intervention, one protein-based intervention, four lipid-based interventions, and seven micronutrient-based interventions (Figure 5A). Improved maternal outcomes included a smaller required insulin dose in mothers with gestational diabetes (following vitamin C supplementation (9)), reduced blood levels of iron and/or zinc (following iron (10) and multiple micronutrient supplementation (11)), decreased total lipid content in placental tissue (following docosahexaenoic acid [DHA] and eicosapentaenoic acid [EPA] supplementation (12)), decreased risk of preeclampsia (following calcium (13), L-arginine and multiple micronutrient supplementation (14)), and decreased risk of placental abruption (following magnesium citrate supplementation (15); Figure 5A).

Of the seven nutritional interventions that associated with improved infant outcomes, two were diet and lifestyle-based, one was protein-based, two were lipid-based, and two were micronutrient-based (Figure 5B). Improved infant outcomes included improved fetal growth (following lipid supplementation (16)), increased fetal nervonic acid ratio (following EPA and DHA supplementation (17)), and a decreased risk of intrauterine growth restriction (IUGR; following L-arginine supplementation (18)), twin-to-twin transfusion syndrome incidence (TTTS [a syndrome that can occur in diamniotic monozygotic twin pregnancies that creates an imbalance in nutrient allocation between fetuses]; following Ensure® liquid supplementation (19)), and preterm birth (for infants born to mothers who smoked; following vitamins C+E (supplementation with both vitamins concurrently) (20); Figure 5B).

***Interventions associated with placental phenotype are more likely, overall, to improve maternal and offspring outcomes***

*Maternal outcomes positively associate with placental changes*

Of the eight micronutrient-based intervention studies that reported improved maternal outcomes, seven (89%) reported associations between the intervention and placental phenotype. Placental changes and improved maternal outcomes included: a reduction in oxidative stress markers in both the placental tissue and maternal blood plasma (following vitamin C supplementation starting once the mother was diagnosed with gestational diabetes (9)) and increased mRNA expression of placental iron uptake transferrin receptor 1 and increased maternal iron and zinc levels (after multiple micronutrient supplementation initiated in the second trimester (11)), and decreased risk of placental abruption (which was classified as both a placental and maternal outcome in this study; following magnesium citrate supplementation starting in the second trimester (15)). Additionally, reduced risk of preeclampsia was reported in four studies, as discussed above (13, 14, 21, 22).

For macronutrient-based interventions, all four of the lipid-based studies that reported improved maternal outcomes also reported placental changes following nutritional intervention. Omega-3 supplementation beginning in the first trimester was associated with increased maternal plasma DHA and EPA levels and decreased expression of placental factors like interleukin 6 and 8 (23), whilst omega-3 supplementation beginning in the second trimester was associated with increased maternal plasma DHA and EPA enrichment, circulating maternal DHA and EPA (73), and decreased total lipid content and ability of the placenta to store and esterify lipids (12). Further, daily fish oil supplementation initiated during the third trimester was associated with increased maternal circulating DHA and plasma nervonic acid content and increased fatty acid concentration in placental tissue (17). Reduced risk of preeclampsia, but not placental abruption, was reported in one protein-based study as discussed above (14).

One of the four diet and lifestyle-based interventions that included maternal outcome data reported improved maternal outcomes following intervention without changes in the placenta (24).

*Offspring outcomes positively associate with placental changes*

Of the 12 micronutrient-based interventions that included data on offspring outcomes, two (17%) reported changes in placental phenotype and improved offspring outcomes following nutritional intervention. In mothers with gestational diabetes mellitus, daily supplementation with vitamin C from diagnosis onwards was associated with a reduction in both fetal and placental oxidative stress markers and newborn birthweight, in comparison to the non-treatment group (9). A decreased risk of both preterm birth and placental abruption was also reported in a cohort of mothers who smoked during pregnancy following daily vitamins C+E supplementation initiated during the first trimester (20). One of the 12 studies with offspring outcome data also reported an adverse offspring outcome (increased incidence of preterm birth) following daily ferrous gluconate supplementation from preconception to birth, despite noting a reduced risk of chorioamnionitis (1).

Of the macronutrient-based interventions, two of four lipid-based interventions that reported offspring outcome data observed altered placental phenotype and improved offspring outcomes. SQLNS supplementation initiated preconceptionally was associated with improved fetal growth, increased placental area (16), and changes in expression of multiple placental metabolic genes. Fish oil supplementation initiated in the third trimester was associated with higher offspring plasma nervonic acid content, increased polyunsaturated placental fatty acid composition, but no changes to visual or cognitive development outcomes measured in the offspring (17). Additionally, two protein-based interventions reported on offspring outcomes, with one noting increased cerebro-placental ratio and decreased incidence of intrauterine growth restriction (IUGR) following daily L-arginine supplementation initiated during the third trimester (18).

Improved offspring outcomes were reported in two diet and lifestyle-based intervention studies, one of which also reported placental changes following the intervention. First, Ensure® liquid supplementation initiated upon diagnosis of a monochorionic diamniotic pregnancy was associated with decreased risk of TTTS and incidence at delivery time (19). Second, daily chlorella supplementation beginning in the second trimester was associated with decreased fetal dioxin transfer (measured through decreased total toxic equivalents), but no significant change in toxin concentrations pre- and post-treatment (25). Notably, we determined the second study to have a high RoB.
