## Supplementary Tables for "Do nutritional interventions before or during pregnancy affect placental phenotype? Findings from a systematic review of human clinical trials"

**Supplementary Table S1.** Synthesis without meta-analysis (SWiM) framework reporting item checklist.

| SWiM reporting item | Item description | Page in manuscript where item is reported |
|---|---|---|
| **1** Grouping studies for synthesis | 1a) Provide a description of, and rationale for, the groups used in the synthesis (e.g., groupings of populations, interventions, outcomes, study design) | 4 |
| | 1b) Detail and provide rationale for any changes made subsequent to the protocol in the groups used in the synthesis | 5 |
| **2** Describe the standardised metric and transformation methods used | Describe the standardised metric for each outcome. Explain why the metric(s) was chosen, and describe any methods used to transform the intervention effects, as reported in the study, to the standardised metric, citing any methodological guidance consulted | N/A – not possible for the varied types and scope of outcomes reported. |
| **3** Describe the synthesis methods | Describe and justify the methods used to synthesise the effects for each outcome when it was not possible to undertake a meta-analysis of effect estimates | 4 |
| **4** Criteria used to prioritise results for summary and synthesis | Where applicable, provide the criteria used, with supporting justification, to select the particular studies, or a particular study, for the main synthesis or to draw conclusions from the synthesis (e.g., based on study design, risk of bias assessments, directness in relation to the review question) | 5-6 |
| **5** Investigation of heterogeneity in reported effects | State the method(s) used to examine heterogeneity in reported effects when it was not possible to undertake a meta-analysis of effect estimates and its extensions to investigate heterogeneity | N/A – studies reported on unique interventions and outcomes |
| **6** Certainty of evidence | Describe the methods used to assess certainty of the synthesis findings | 5-7 – Risk of bias assessment was performed, though synthesis for specific intervention types & outcomes was not possible, given the vast assortment and heterogeneity of outcomes reported |
| **7** Data presentation methods | Describe the graphical and tabular methods used to present the effects (e.g., tables, forest plots, harvest plots). | 6 |

| | | |
|---|---|---|
| | Specify key study characteristics (e.g., study design, risk of bias) used to order the studies, in the text and any tables or graphs, clearly referencing the studies included | |
| **8** Reporting results | For each comparison and outcome, provide a description of the synthesised findings, and the certainty of the findings. Describe the result in language that is consistent with the question the synthesis addresses, and indicate which studies contribute to the synthesis | 6-13 – certainty of findings was assessed according to risk of bias assessment |
| ***Discussion*** | | |
| **9** Limitations of the synthesis | Report the limitations of the synthesis methods used and/or the groupings used in the synthesis, and how these affect the conclusions that can be drawn in relation to the original review question | 14-15 |

**Supplementary Table S2.** Categories of placental measures for the purpose of this review.

| Placental measure | Definition | Examples (non-exhaustive) |
|---|---|---|
| Anthropometry | Any physical characteristics related to the placenta | Weight, cerebro-placental ratio, fetal-placental ratio, area |
| Molecular | Any alterations to placental genotype, gene expression, or molecular changes | Placental docosahexaenoic acid content, fatty acid synthase expression |
| Pathology | Any pathological findings related to the placenta performed by a pathologist | Chorioamnionitis, microscopic placental malaria risk |
| Placental abruption | Cases of placental abruption | Placental abruption |
| Placenta-related disease | Any disease with placental origins, or that significantly impacts the placenta | Preeclampsia, premature rupture of membranes, twin-to-twin transfusion syndrome, placental malaria |

**Supplementary Table S3.** Cochrane Collaboration's Tool for Assessing Risk of Bias checklist of assigned criterion.

| Checklist item requiring specification | Assigned criterion |
|---|---|
| Random sequence generation | 1. Was the randomization sequence or method reported? |
| Allocation concealment | 1. Was the allocation sequence and participant assignment concealed from parties directly involved (e.g. offsite data manager controlled the randomization assignment)? |
| Blinding of participants and personnel | 1. Were both participants and personnel blinded to which treatment was being received? |
| Blinding of outcome assessment | 1. Was the outcome objective (lower risk of bias) or subjective (higher risk of bias)? |
| Incomplete outcome data | 1. Was placental data provided for each member of the cohort? Or just a selected group?<br>2. If the entire cohort, were attrition rates and acceptability low or high?<br>3. If a subset of the group was selected, how was this subset created? |
| Selective outcome reporting | 1. Are both significant and non-significant results reported?<br>2. Do initial plans for placental data reporting match what was actually reported? |
| Other (uptake of interventions/intervention adherence) | 1. Was uptake and adherence measured and reported by authors?<br>2. How often did researchers follow-up with participants? |

**Supplementary Table S4.** Maternal comorbidities selected for or noted in included studies.

| **Maternal comorbidity*** | **Number of studies** |
|---|---|
| None | 20 |
| Preeclampsia | 12 |
| Obesity/Overweight | 7 |
| Hypertension | 7 |
| Diabetes | 6 |
| Smoking | 6 |
| Anemia | 3 |
| Nutritional deficiency | 3 |
| HIV | 2 |
| Allergy status | 1 |

*Some studies selected or noted more than one maternal comorbidity.

**Supplementary Table S5**. Summary of studies with primary outcomes relevant to maternal health.

| First author, year (citation) | Key findings related to maternal health | Significantly altered maternal outcome?* | Significant placental phenotype alterations?* |
|---|---|---|---|
| He, 2020 (1) | No difference to chorioamnionitis rate | No | Yes |
| de Araújo, 2020 (2) | No difference in preeclampsia or eclampsia, gestational hypertension, maternal stroke, death. Decreased risk of placental abruption (defined as a maternal composite in this study). | Yes | Yes |
| Zheng, 2020 (3) | Lower rates of preeclampsia in high dose group | Yes | Yes |
| Hofmeyr, 2019 (4) | No difference in preeclampsia risk | No | No |
| Ormesher, 2018 (5) | No significant reduction in blood pressure compared to placebo | No | No |
| Wen, 2018 (6) | No significant differences in preeclampsia risk | No | No |
| Lager, 2017 (7) | No significant difference to maternal inflammatory status, insulin sensitivity, or circulating lipids | No | Yes |
| Bujold, 2017 (8) | No significant difference in uterine artery pulse index compared to low-dose group | No | No |
| Jobarteh, 2017 (9) | Increased maternal iron and zinc status | Yes | Yes |
| Devi, 2017 (10) | Increased homocysteine remethylation in late pregnancy | Yes | No |
| Maged, 2016 (11) | Supplementation significantly improved oxidative stress markers (GSH, MDA, SOD, CAT, GPx), improved maternal blood sugar control | Yes | Yes |
| Calabuig-Navarro, 2016 (12) | Improved maternal inflammatory status; increased maternal plasma n3:n6 ratio, increased maternal plasma enrichment of DHA and EPA | Yes | Yes |
| Abramovici, 2015 (13) | No difference in preeclampsia risk | No | Yes |
| Etheredge, 2015 (14) | Significantly improved hemoglobin and iron status. Anemia risk reduced by 40%, risk of iron deficient anemia by 66%. | Yes | No |
| Hurtado, 2015 (15) | Increased DHA levels and nervonic acid ratio. | Yes | Yes |
| Haghiac, 2015 (16) | Improved maternal inflammatory status via increased plasma DHA and EPA, and n-3 FA:n-6 FA ratio higher in treatment group. | Yes | Yes |
| Keelan, 2015 (17) | Increased maternal circulating DHA and EPA levels | Yes | Yes |
| Kiondo, 2014 (18) | No significant differences in preeclampsia risk | No | No |

| | | | |
|---|---|---|---|
| Milman, 2014 (19) | No significant difference in iron deficiency or iron deficiency anemia between treatment groups | No | No |
| Parrish, 2013 (20) | No significant differences in preeclampsia risk | No | No |
| Johnston, 2013 (21) | No significant differences in preeclampsia risk | No | No |
| Jiang, 2013 (22) | Decreased placental sFLT1 production, which is an antiangiogenic factor linked with preeclampsia | Yes | Yes |
| Zhou, 2012 (23) | No significant differences in preeclampsia or gestational diabetes mellitus risk | No | No |
| Vadillo-Ortega, 2011 (24) | Decreased risk of preeclampsia | Yes | Yes |
| McCance, 2010 (25) | No significant differences in preeclampsia risk | No | No |
| Roberts, 2010 (26) | No significant difference pregnancy-associated hypertension, thrombocytopenia, elevated serum creatinine levels, or eclamptic seizure | No | No |
| Villar, 2009 (27) | No significant differences in preeclampsia risk | No | No |
| Spinnato, 2007 (28) | Increased risk of premature rupture of membranes (PROM), no significant differences in preeclampsia risk | Yes** | Yes |
| Villar, 2006 (29) | Decreased severity of preeclampsia by 35 weeks' gestation, eclampsia, and hypertension | Yes | Yes |
| Rumbold, 2006 (30) | No significant differences in preeclampsia risk | No | No |

*Significance was determined by the statistical tests performed in each study.
**Alterations to maternal outcomes were adverse effects.

**Supplementary Table S6**. Summary of studies with primary outcomes relevant to offspring health.

| First author, year (citation) | Key findings related to offspring health | Significantly altered offspring outcome?* | Significant placental phenotype alterations?* |
|---|---|---|---|
| Castillo-Castrejon, 2021 (31) | Improved fetal growth due to activated mTOR and IGF-1 signalling in Pakistan cohort | Yes | Yes |
| He, 2020 (1) | No difference in childhood asthma rates | No | Yes |
| de Araújo, 2020 (2) | No difference in rates of PT birth, stillbirth, neonatal death, NICU admission, or low birthweight | No | Yes |
| Brabin, 2019 (32) | Increased preterm birth risk in treatment arm, no difference in growth restriction | Yes** | Yes |
| Kashanian, 2018 (33) | No difference in preterm rupture of membrane (PROM) or premature preterm rupture of membrane (PPROM) risk | No | No |
| Maged, 2016 (11) | Decreased fetal MDA, increased SOD. Neonatal birth weight significantly lower in treatment group (3.267kg vs 3.98kg) | Yes | Yes |
| Abramovici, 2015 (13) | Decreased risk of preterm birth in smokers group | Yes | Yes |
| Gernand, 2015 (34) | No difference in intrauterine growth factors or birth weight | No | No |
| Etheredge, 2015 (14) | No difference in birth weight | No | No |
| Hurtado, 2015 (15) | Increased nervonic acid ratio in plasma and erythrocyte lipids. No difference in visual, cognitive, or psychomotor development. | Yes | Yes |
| Wietrak, 2015 (35) | No difference to gestational length, birth weight, or Apgar scores | No | Yes |
| Roberts, 2010 (26) | No difference in preterm birth, fetal-growth restriction, or perinatal death risk | No | No |
| Winer, 2009 (36) | No difference in birth weight | No | No |
| Villar, 2009 (27) | No difference in low birth weight, small for gestational age, or perinatal death risk | No | Yes |
| Chiossi, 2008 (37) | Decreased incidence of twin-to-twin transfusion syndrome (TTTS) and prevalence of TTTS at delivery. Increased time between TTTS diagnosis and delivery. | Yes | Yes |
| Rumbold, 2006 (30) | No difference in fetal death or small for gestational age risk. | No | No |
| Rytlewski, 2006 (38) | Decreased risk of intrauterine growth restriction (IUGR). No difference in fetal death risk, birth weight, or gestational length. | Yes | Yes |

| | | | |
|---|---|---|---|
| Villar, 2006 (29) | No difference in preterm birth risk | No | No |
| Nakano, 2005 (39) | Decreased maternal total toxic equivalents (TEQ), meaning decreased maternal dioxin transfer to child | Yes | No |
| Helland, 2001 (40) | No difference in gestational length or birth weight | No | No |

*Significance was determined by the statistical tests performed in each study.
**Alterations to infant/fetal outcomes were adverse effects.

**Supplementary Table S7.** Summaries of studies with relevant key placental findings.

| First author, year (citation) | Location | Number of groups (group types); n | Treatment intervention | Composition (daily, unless noted otherwise) | Key placental findings following treatment | Significant placental phenotype alterations?* |
|---|---|---|---|---|---|---|
| Castillo-Castrejon, 2021 (31) | Guatemala and Pakistan | 4 (1 control and treatment per location); n=12,12,12,12 | SQLNS micronutrient supplement | linoleic 4.9 g and α-linolenic 0.59 g | -Average placental area ($cm^2$) larger in treatment groups<br>-In Pakistani cohort, increased rpS6(T37/46):rpS6 (ribosomal protein S6) ratio 1.5 fold, decreased AMPKA(T172):AMPKA (AMP-activated protein kinase) ratio<br>-Increased pregnancy-associated plasma protein A (PAPP-A) in both treatment cohorts | Yes |
| Awe, 2020 (41) | United States of America (USA) | 1, split into D3 sufficient and deficient after treatment (treatment); n=43 | Vitamin D3 + standard prenatal vitamin | 4000IU D3 + 400IU from prenatal | -Near delivery, lower placental Fms Related Receptor Tyrosine Kinase 1 (Flt-1), SRC associated in mitosis of 68kDA (Sam68) than Vitamin D3 deficient placentas<br>-No difference in sFLT-1 | Yes |
| He, 2020 (1) | USA | 2 (<32 ng/mL serum hydro cholecalciferol status, >32 ng/mL); n=27,20 | Vitamin D | 400 vs 4000 IU Vitamin D | -No difference in placental abruption risk<br>- Lower Integrator Complex Subunit 9 (INTS9), von Willebrand factor (vWF), Metastasis Associated in Colon Cancer-1 (MACC1), Age-related Maculopathy Susceptibility 2 (ARMS2) expression<br>-Increased contactin 5 (CNTN5) gene expression, chorionic villi surface density in higher concentration group | Yes |
| de Araújo, 2020 (2) | Brazil | 2 (placebo, treatment); n=422,407 | Magnesium citrate | 300 mg magnesium citrate | -Lower placental abruption risk (2.2% vs. 5.0%) | Yes |

| | | | | | | |
|---|---|---|---|---|---|---|
| Zheng, 2020 (3) | China | 2 (low dose, high dose); n=378,410 | Folic Acid | 0.4mg FA, 4mg FA | -Lower preeclampsia risk with high treatment compliance<br>-No difference in placental abruption risk | Yes |
| Brabin, 2019 (32) | Burkina Faso | 2 (control, treatment); n=144,163 | Iron + folic acid | 60mg ferrous gluconate + 2.8mg folic acid | -Reduced grade 2 and 3 chorioamnionitis risk (40.4% vs 48.9%)<br>-No difference in placental malaria risk | Yes |
| Hofmeyr, 2019 (4) | South Africa, Zimbabwe, Argentina | 2 (placebo, treatment); n=283,298 | Calcium | 500 mg calcium | -No difference in placental abruption risk | No |
| Ormesher, 2018 (5) | United Kingdom (UK) | 2 (placebo, treatment); n=21,20 | Beetroot juice | ~400mg nitrate in 70mL beetroot juice | -No difference in uteroplacental blood flow | No |
| Wen, 2018 (6) | Argentina, Australia, Canada, Jamaica, UK | 2 (placebo, treatment); n=1179,1172 | Folic acid | 4.0mg from 8-16 weeks, 1.1mg until birth | -No difference in placental abruption risk | No |
| Kashanian, 2018 (33) | Iran | 2 (placebo, treatment); n=111, 127 | Copper | 1000mg copper | -No difference in placental abruption or placenta previa risk | No |
| Lager, 2017 (7) | USA | 2 (placebo, treatment); n=19,19 | DHA | DHA 800mg w/ algal oil, placebo was corn/soy oil. | -Increased placental membrane DHA levels, fatty acid transporting protein 4 expression<br>-Decreased placental inflammation via decreased nuclear factor of kappa light polypeptide gene enhancer in B-cells inhibitor alpha (IkBa), p38 mitogen-activated protein kinase alpha (p38α), and increased cyclo-oxygenase 2 (COX-2) and nuclear factor kappa B p65 activation (NF-kB)<br>-Decreased fetal-placental ratio<br>-No difference in placental weight | Yes |

| | | | | | | |
|---|---|---|---|---|---|---|
| Park, 2017 (42) | USA | 1 (treatment); n=24 | Vitamin D | 511IU (311IU from diet, 200IU via multivitamin ) | -Increased LDL Receptor Related Protein 2 (LRP2), cubulin (CUBN), Cytochrome P450 Family 2 Subfamily R Member 1 (CYP2R1), Cytochrome P450 Family 24 Subfamily A Member 1 (CYP24A1), Cytochrome P450 Family 27 Subfamily B Member 1 abundance (CYP27B1) | Yes |
| Bujold, 2017 (8) | Canada | 2 (low dose, high dose); n=131 total | Low or high dose flavanol and theobromine chocolate | 69mg Theobromine, 25.4mg Epicatechins, 6.3mg Catechins, 32.1mg Dimers, 107.6mg Trimers-decamers | -No difference in preeclampsia risk via uterine artery pulsatility index | No |
| Jobarteh, 2017 (9) | Gambia | 4 (iron + folate (FeFol), MMN, PEB, PEB+MMN); n=74,76,76,75 | Multi-micronutrient (MMN) and/or Protein-Energy (PEB) Ball | FeFol: 60mg/d, 400 ug folic acid. MMNs: combination of 15 micronutrients. Pe: same as FeFol plus energy, protein, lipids. PE + MMNs: MMNs content plus PE content. | -Placental iron uptake protein transferrin receptor 1 mRNA levels 30-49% higher in PE and PE+MMN arms than FeFol arm. 29% higher in PE+MMN arm than just MMN arm.<br>-No difference in placental weight, breadth, or length | Yes |

| | | | | | | |
|---|---|---|---|---|---|---|
| Devi, 2017 (10) | India | 3 (Placebo, placebo+milk, milk+ B12); n=31,29,29 | Vitamin B-12 | 500 mL/d milk, and/or 100ug vitamin b-12 tablet/d | -No difference in placental mRNA for methionine pathways, placental long interspersed nuclear elements 1 (LINE-1), and vascular endothelial growth factor (VEGF) promoter methylation. | No |
| Darling, 2017 (43) | Tanzania | 4 (placebo, VitA, Zinc, VitA+Zinc); n=611,613,608, 602 | Vitamin A and/or Zinc | 2500 IU VitA, and/or 25mg zinc | -36% lower risk of histopathological-positive placental malaria result with zinc supplementation | Yes |
| Maged, 2016 (11) | Egypt | 2 (control, treatment); n=100,100 | Vitamin C | 1 gram L-ascorbic acid | -Lower glutathione (GSH), malondialdehyde (MDA), catalase (CAT), glutathione peroxidase (GPx)<br>-Higher superoxide dismutase (SOD) | Yes |
| Calabuig-Navarro, 2016 (12) | USA | 2 (placebo, treatment); n=16,17 | LC-PUFA | 2000 mg n-3LCPUFA (800mg DHA + 1200mg EPA) | -Lower fatty acid synthase (FAS), diacylglycerol O-acyltransferase 1 (DGAT1), peroxisome proliferator–activated receptor γ (PPAR), perilipin 2 (PLIN2) | Yes |
| Johnston, 2016 (44) | Ireland | 2 (placebo, treatment); n=30,27 | Vitamin C + E | 1000mg vitamin C, 400IU Vitamin E | -No difference in placental antioxidant enzymes and lipid peroxidation | No |
| Owens, 2015 (45) | Gambia | 2 (placebo, treatment); n=247,239 | Multi-micronutrient (UNIMMAP) | Vitamin A (800 retinol equivalents), D (200 IU), E (10 mg), C (70 mg), thiamin (1.4 mg), riboflavin (1.4 mg), niacin (18 mg), | -No difference in plasminogen activator inhibitor-1 (PAI-1), plasminogen activator inhibitor-2 (PAI-2), PAI-1:PAI-2 ratio<br>-No difference in placental weight | No |

| | | | | | | |
|---|---|---|---|---|---|---|
| | | | | pyridoxine (1.9 mg), cobalamin (2.6 mg), folic acid (400 mg), iron (30 mg), zinc (15 mg), copper (2 mg), selenium (65 mg), iodine (150 mg) | | |
| Abramovici, 2015 (13) | USA | 4 (smoker placebo, smoker treatment, non-smoker placebo, non-smoker treatment); n=763,788,4213,4205 | Vitamin C + E | 1000 mg Vitamin C, 400 IU vitamin E | -Reduced placental abruption risk in smoker cohorts (1.5% vs 0.1%)<br>-No difference in preeclampsia risk | Yes |
| Gernand, 2015 (34) | Bangladesh | 2 (Iron + folic acid (IFA), MM); n=191,205 | Multi-micronutrient | IFA: 27mg iron + 600ug FA<br>MM: IFA + vitamins A (770 µg retinol equivalents), D (5 µg), E (15 mg), B12(2.5 mg), B6 (1.9 mg), C (85 mg), thiamin (1.4 mg), | -No difference in placental weight | No |

| | | | | | | |
|---|---|---|---|---|---|---|
| | | | | riboflavin (1.4 mg), niacin (1.4 mg), zinc (12 mg), iodine (220 µg), copper (1000 µg), selenium (60 µg) | | |
| Etheredge, 2015 (14) | Tanzania | 2 (placebo, treatment); n=510,493 | Iron | 60 mg iron | -No difference in weight, or microscopic and submicroscopic placental malaria risk | |
| Hurtado, 2015 (15) | Spain | 2 (control, treatment); n=38,38 | Fish oil | 18 mg/100mL EPA, 80 mg/mL DHA | -Higher placental fatty acid composition in phospholipids of elaidic acid (C18:ln-9), linoleic acid (C18:2n-6), docosahexaenoic acid (DHA), omega-3 polyunsaturated fatty acid (n-3 PUFA)<br>-No difference to alpha-linolenic acid (C18:3n-3), 15-tetracosenoic acid (C24:1n-9), eicosapentaenoic acid (EPA), saturated fatty acid (SFA), monounsaturated fatty acid (MUFA), omega-6 polyunsaturated fatty acid (n-6 PUFA) | Yes |
| Haghiac, 2015 (16) | USA | 2 (placebo, treatment); n=25,24 | Omega-3 fatty acid supplement | 800mg DHA and 1200mg EPA | -Lower placental interleukin 6 and 8 (IL6, IL8) tumour necrosis factor alpha (TNFα), toll-like receptor 4 (TLR4) mRNA expression<br>-No difference to total omega-3 fatty acid (n-3 PUFA) concentration | Yes |
| Keelan, 2015 (17) | Australia | 2 (placebo, treatment); n=28,22 | Omega-3 polyunsaturated fatty acids (n-3 PUFAs) | 4 pills, 3.7g n-3 PUFAS (56% DHA, 27.7% EPA) per tablet | -Increased placental DHA (80%), TNFα (14x), 18-hydroxyeicosapentaenoic acid (18-HEPE) | Yes |

| | | | | | | |
|---|---|---|---|---|---|---|
| | | | | | -No difference to interleukin 1beta, 6, or 10 (IL1B, IL6, IL10), Prostaglandin-Endoperoxide Synthase 2 (PTGS2) | |
| Wietrak, 2015 (35) | Poland | 2 (control, treatment); n=50,28 | DHA | 300 mg DHA | -Lower average placental weight (510g vs 530g)<br>-No difference in protein expression of cyclin-dependent kinase inhibitor 1 (p21) or antigen Ki-67 | Yes |
| Kiondo, 2014 (18) | Uganda | 2 (placebo, treatment); n=418,415 | Vitamin C | 1000mg Vitamin C | -No difference in preeclampsia incidence or placental abruption risk | No |
| Milman, 2014 (19) | New Zealand | 2 (ferrous bisglycinate, ferrous sulfate); n=40,40 | Iron | 25mg ferrous bisglycinate elemental iron, or 50mg ferrous sulfate elemental iron | -No difference in immunostaining for transferrin receptor | No |
| Parrish, 2013 (20) | USA | 2 (placebo, treatment); n=135,132 | Phytonutrient | ~7.5mg beta-carotene, 234mg vitamin C, 30mg vitamin E, 420mg folate, 60mg calcium | -No difference in placental abruption or PPROM risk | No |
| Johnston, 2013 (21) | Ireland | 2 (placebo, treatment); n=382,379 | Vitamin C + E | 1000mg Vitamin C, 400 IU vitamin E | -No difference in placental abruption risk | No |
| Jiang, 2013 (22) | USA | 2 (low dose, high dose); n=12,12 | Choline | 480mg/d or 930mg/d choline through | -30% downregulation of soluble fms-like tyrosine kinase-1 (sFLT1)<br>-43 placental genes upregulated, of note ghrelin and obestatin prepropeptide | Yes |

| | | | | | | |
|---|---|---|---|---|---|---|
| | | | | 380mg/d from diet, plus either 100 or 550mg from supplemental choline chloride. | (GHRL; 1.6x increase), neuropeptide Y (NPY) receptor 75 (1.7x increase), elastin (2.03x increase)<br>-123 genes downregulated<br>-Treatment alters 197 biological processes in higher dose | |
| Jiang, 2012 (46) | USA | 2 (low dose, high dose); n=12,12 | Choline | 480mg/d or 930mg/d choline through 380mg/d from diet, plus either 100 or 550mg from supplemental choline chloride. | -Increased placental promoter methylation of cortisol-regulating genes, corticotropin releasing hormone (CRH) and glucocorticoid receptor (NR3C1)<br>-Increased placental global DNA methylation and demethylated histone H3 at H3K9me2<br>-Lower placental corticotropin-releasing hormone (CRH) transcript abundance | Yes |
| Zhou, 2012 (23) | Australia | 2 (control, treatment); n=1202,1197 | DHA fish oil | 3 pills, 500mg each totalling to 800mg DHA | -No difference in placental weight or preeclampsia risk | No |
| Vadillo-Ortega, 2011 (24) | Mexico | 3 (placebo, L-arg+vitamins, vitamins); n=222,228,222 | L-arginine (L-arg) and/or vitamins | L-arg bar: 6.6 g L-arg, plus vitamins from vitamin-only bar (listed next) from 2 bars. Vitamins: no L-arg but other vitamins: C | -Lower preeclampsia risk (30% in placebo vs 13% in L-arg + vitamin group vs 23% in vitamin group)<br>-No difference in placental abruption risk | Yes |

| | | | | | | |
|---|---|---|---|---|---|---|
| | | | | (250mg), B6 (2.0mg), B12 (4.8ug), E (200IU), niacin (25mg), folate (200ug). | | |
| McCance, 2010 (25) | Ireland, Scotland, England | 2 (placebo, treatment); n=382,379 | Vitamin C + E | 1000mg Vitamin C, 400 IU vitamin E | -No difference in placental abruption or preeclampsia risk | No |
| Mercer, 2010 (47) | USA | 2 (placebo, treatment); n=39,34 | Vitamin C + E | 500mg Vitamin C, 200IU vitamin E | -No difference in amnio-choriodecidua separation risk | No |
| Roberts, 2010 (26) | USA | 2 (placebo, treatment); n=4976,4993 | Vitamin C + E | 1000mg Vitamin C, 400IU Vitamin E | -No difference in placental abruption or preeclampsia risk | No |
| Winer, 2009 (36) | France | 2 (placebo, treatment); n=22,21 | L-arginine | 14g (90cc) ARG | -No difference in placental weight, placental abruption, hypotrophy, thrombosis, or histological abnormality risk | No |
| Villar, 2009 (27) | India, Peru, South Africa, Vietnam | 2 (placebo, treatment); n=678,687 | Vitamin C + E | 1000mg Vitamin C, 400 IU vitamin E | -No difference in placental abruption or preeclampsia risk | No |
| Chiossi, 2008 (37) | USA | 2 (standard diet, treatment); n=51,52 | Ensure liquid nutritional supplement | 250kcal, 6g lipids, 40g carbohydrates, 9g protein, vitamins, minerals | -Decreased risk of Twin-to-twin transfusion syndrome (TTTS) diagnosis (15.5% vs 38%), and TTTS at delivery (11.8% vs 34.6%) | Yes |

| | | | | | | |
|---|---|---|---|---|---|---|
| Spinnato, 2007 (28) | Brazil | 2 (placebo, treatment); n=349,351 | Vitamin C + E | 1000mg Vitamin C, 400IU vitamin E | -Increased risk of PROM (5.5% vs 10.6%)<br>-No difference in placental abruption risk | Yes |
| Kaplas, 2007 (48) | Finland | 3 (control/placebo, diet/placebo, diet/probiotics); n=8,12,10 | Diet and/or probiotics | Diet: Overall monounsaturated fatty acids (MUFA) contributing 10–15%, PUFA 5–10%, and saturated fatty acids (SFA) 10% or less of energy in-take. Probiotics: capsules containing Lactobacillus rhamnosus GG and Bifidobacterium lactis Bb12. Exact composition not listed. | -Increased n-3 PUFA, Dihomo-c-linolenic acid (DHGA), and eicosatetraenoic acid (20:4n-3) in diet group<br><br>-Increased DHGA, EPA, arachidonic acid, linoleic acid (LA), 20:4n-3 in probiotic group. Changes to DHGA and linoleic acid attributed to probiotics only. | Yes |

| | | | | | | |
|---|---|---|---|---|---|---|
| van Eijk, 2007 (49) | Kenya | 3 groups each separated into 2 for HIV+ or HIV- status (0 intervention, 1, 2); n=1172,1140,796 | Haematinics and/or IPTp-SP | Haematinics: 200 mg ferrous sulphate + 5mg folic acid. IPTp-SP: intermittent preventative treatment with sulphadoxine-pyrimethamine | -Decreased placental malaria risk for group using 2 interventions compared to reference group of no intervention (0.56 overall, 0.43 for HIV+, 0.61 for HIV-) | Yes |
| Larqué, 2006 (50) | Spain | 4 (placebo, methyltetrahydro-folate (MTHF), DHA, DHA+MTHF); n=26,27,25,21 | MTHF and/or DHA | MTHF: 400ug 5-methyl-tetrahydrofolic acid, DHA: 500mg DHA + 150mg eicosapentaenoic acid | -Increased DHA in placental phospholipids at delivery, proportion of DHA in placental phospholipids and mRNA expression in the membrane proteins fatty acid transport proteins 1 and 4 (FATP-1, FATP-4)<br>-No difference in placental tissue mRNA expression of FATP-1, FATP-4, FATP-6, fatty acid translocase (FAT/CD36), plasma membrane fatty acid–binding protein (FABPpm/GOT2), heart fatty-acid transport protein (H-FABP), and arachidonic acid (AA) proportion<br>-No difference in placental weight | Yes |
| Rytlewski, 2006 (38) | Poland | 2 (placebo, treatment); n=31,30 | L-arginine | 3g L-arginine (6*0.5g tablets) | -Increased cerebro-placental ratio after 4 weeks (1.5 to 1.71 vs 1.41 to 1.09) | Yes |

| | | | | | | |
|---|---|---|---|---|---|---|
| Klingler, 2006 (51) | Spain | 4 (placebo, fish oil+5-MTHF, fish oil, 5-MTHF); n=12,11,16,16 | Modified fish oil and/or MTHF | Fish oil: 500mg DHA + 150mg EPA. MTHF: 400ug 5-MTHF (delivered as 800ug 6,RS,5-methyltetrahydrofolate) | -Proportion of DHA in placental phospholipids similar between fish oil groups, but higher than non-fish oil groups<br>-Increased proliferation cell nuclear antigen (PCNA) in placentas of fish oil + 5-MTHF group than placebo by 66%<br>-Increased EPA in fish oil groups<br>-No difference in placental p53 or cytokeratin levels | Yes |
| Rumbold, 2006 (30) | Australia | 2 (placebo, treatment); n=942,935 | Vitamin C + E | 1000mg Vitamin C + 400 IU Vitamin E (from a combined 4 pills) | -No difference in placental abruption or preeclampsia risk | No |
| Villar, 2006 (29) | Argentina, Egypt, South Africa | 2 (placebo, treatment); n=4161,4151 | Calcium | 1.5g calcium | -Decreased risk of preeclampsia by 35 weeks gestation (1.2% vs 2.8%)<br>-No difference in placental abruption risk | Yes |
| Cox, 2005 (52) | Ghana | 2 (placebo, treatment); n=38,38 | Vitamin A | 10,000IU vitamin A as retinyl palmitate | -No difference in active or chronic-active placental malaria risk in current pregnancy compared to past resolved infection at delivery | No |
| Nakano, 2005 (39) | Japan | 2 (control, treatment); n=21, 23 | Chlorella | 6g chlorella (30 tablets/day, 10 after each meal), composed of (in g/100g): moisture, 4.7; | -No difference in pg/whole sample g for polychlorinated dibenzo-p-dioxins (PCDD), poly-chlorinated dibenzofurans (PCDF), co-planar polychlorinated biphenyls (Co-PCB), total toxic equivalents (TEQ) | No |

| | | | | | | |
|---|---|---|---|---|---|---|
| | | | | chlorophyll, 2.4; dietary fiber, 9.8; protein (N · 6.5), 57.8; lipid, 10.4. | | |
| Pressman, 2003 (53) | USA | 2 (control, treatment); n=10,10 | Prenatal vitamin plus Vitamin C + E | Prenatal (120mg Vitamin C + 30 IU vitamin E) + 400 IU vitamin E + 500 mg vitamin C | -No difference in chorioamnion grams to burst (gf), chorioamnion maximal deflection (mm), or chorioamnion Vitamin E levels | No |
| Helland, 2001 (40) | Norway | 2 (placebo, treatment); n=166,175 | Cod liver oil | 10mL cod oil (117ug/mL vitamin A, 1 ug/mL Vitamin D, 1.4 mg/mL dl-alpha-tocopherol | -No difference in placental weight | No |

*Significance was determined by the statistical tests performed in each study.
